# Comparing screening frameworks for populations with multiple overlapping high-risk factors: A case of tuberculosis screening in China

**DOI:** 10.64898/2026.07.08.26357439

**Authors:** Wenyong Zhou, Zexuan Wen, Tao Li, Xiaoqiu Liu, Canyou Zhang, Yunzhou Ruan, Hui Zhang, Nimalan Arinaminpathy, Weibing Wang

**Affiliations:** Shanghai Institute of Infectious Disease and Biosecurity, School of Public Health, Fudan University, Shanghai 200032, China; Department of Epidemiology, School of Public Health, Fudan University, Shanghai 200032, China; National Center for Tuberculosis Control and Prevention, Chinese Center for Disease Control and Prevention, Beijing 102206, China; National Key Laboratory of Intelligent Tracking and Forecasting for Infectious Diseases, Chinese Center for Disease Control and Prevention, Beijing, 102206, China; Department of Infectious Disease Epidemiology, School of Public Health, Imperial College London, London W12 0BZ; Department for HIV, Tuberculosis, Hepatitis and STIs, World Health Organisation, Geneva, Switzerland; Key Laboratory of Public Health Safety of Ministry of Education, Fudan University, Shanghai 200032, China; Key Laboratory of Health Technology Assessment, National Health and Family Planning Commission of the People’s Republic of China, Fudan University, Shanghai 200032, China

**Author notes:** Corresponding author: Dr. Weibing Wang, Department of Epidemiology, School of Public Health, Fudan University, 138 Yi Xue Yuan Road, Shanghai 200032, China. These authors contributed equally to this work (Wenyong Zhou, Zexuan Wen). Dr. Nimalan Arinaminpathy and Dr. Weibing Wang are listed as the senior authors of this work.

## Abstract

**Background:** Public health initiatives increasingly target multiple overlapping high-risk groups to maximize impact. However, a common challenge in modelling these initiatives is to capture these overlapping risk factors, leading to potential misallocation of resources and biased effectiveness estimates. Using China’s tuberculosis (TB) control program as an example, this study explores different possible frameworks to account for population heterogeneity and risk overlap.

**Methods:** We examined four risk allocation frameworks: (*i*) Direct Summation (DS), a simple additive benchmark; (*ii*) Probabilistic Union Deduplication (PUD), using inclusion-exclusion principles; (*iii*) Risk population combination (RPC), modeling interaction effects; and (iv) Agent-Based Framework (ABF), a granular microsimulation. To show how these frameworks could be used in epidemiological modelling, we embedded each within a deterministic transmission model of TB epidemiology in China, to simulate the impact of China’s National Tuberculosis Strategic Plan (NTSP). We explored each framework when implemented in both static and dynamic versions. We compared them using methodological principles and indicators of intervention cost (screening volume) and benefits (cases/deaths averted).

**Results:** Under the static version, the detection yield of active cases followed a consistent hierarchy: DS > PUD > RPC ≈ ABF. The DS method systematically overestimated yields by double-counting overlapping populations, while PUD corrected for overlap but ignored interaction. The RPC and ABF methods provided the most granular estimates by incorporating Risk population combinations. Additionally, comparing static versus dynamic versions revealed that for the same multi-risk screening framework, mortality reductions remained stable and incidence reductions varied significantly.

**Conclusion:** This study presents potential screening frameworks for overlapping risk populations. The RPC method offers optimal balance of real-world plausibility and computational efficiency. We propose the dynamic RPC method as the preferred tool for routine analysis where multimorbidity and intersectional risks exist, providing a robust evidence base for optimizing resource allocation in heterogeneous populations.

## Introduction

Screening is the cornerstone of public health programs, aiming to reduce the overall burden of communicable and non-communicable diseases by identifying individuals at high-risk and implementing targeted interventions ^1^. Compared to universal screening strategies, screening of high-risk populations results in fewer adverse effects and is more cost-effective and thus has broad applications, such as tuberculosis (TB), colorectal cancer, lung cancer and so on ^2, 3, 4, 5, 6^. To optimize effectiveness, public health initiatives often require targeting multiple high-risk groups ^7^.

To eliminate TB, the Government of China has proposed the National TB Prevention and Control Plan (2024–2030) ^8^, which subsequently evolved into the National Tuberculosis Strategic Plan (2025–2035) (hereinafter referred to as the NTSP). This Plan includes enhanced active screening measures for TB among high-risk populations (including close contacts, individuals living with HIV, recent TB recovers, the elderly, and individuals living with diabetes mellitus), aiming to identify the most effective integrated interventions and optimize their benefits. Mathematical modelling, an important method for simulating the effectiveness of specific interventions ^9, 10^, will play a pivotal role in evaluating the benefits of this strategy. The key dynamic involves the transfer of individuals between compartments. For instance, when modeling the effect of active case-finding interventions, detected active TB cases should be moved from the active cases compartment into the isolation and treatment compartment. This effectively removes them from the chain of transmission ^11^.

However, a significant challenge is that individuals may belong to multiple high-risk categories simultaneously. For instance, an elderly individual with poorly controlled diabetes may also be a close contact of a TB case. Such overlapping risks derive from complex biological factors (e.g., immunosuppression, comorbidities), behavioral patterns (e.g., smoking, alcohol abuse), or social-environmental exposures (e.g., overcrowded housing, close contact with infectious cases), particularly prevalent in settings with high disease burden and adverse socioeconomic conditions ^12^. Moreover, these risk factors can cluster within individuals, creating challenges beyond simple duplicate counting. A fundamental obstacle for models seeking to address overlapping risks lies not in methodological technique, but in the limited data on both the joint frequency of overlapping risk factors and the nature of their combined effects on TB disease risk. In practice, most screening program evaluation frameworks assess single interventions ^13, 14, 15, 16^, avoiding evaluating the combined effects of multiple interventions^17^, or simply add the outcomes from individual intervention groups ^18^. These approaches may lead to overestimation of target population size and potential effectiveness, resulting in misallocation of public health resources ^19^. Furthermore, most existing studies directly apply community screening coverage rates ^20, 21^. While this approach is easier to model, it is difficult to implement in practice and inconsistent with health economics principles, making it challenging to conduct refined health economic evaluations. To help address this challenge, this study examines different approaches for modelling screening in populations with multiple overlapping high-risk factors, using China’s NTSP-based TB transmission model as a specific case study. We acknowledge that these frameworks do not resolve the underlying data gaps regarding the joint frequency and combined effects of overlapping risk factors; rather, they offer a transparent methodological comparison under explicit assumptions of independently distributed risk factors and multiplicative combined effects. While this application focuses on TB, the proposed methodology can be extended to other public health screening interventions involving overlapping high-risk groups.

## Methods

### Study design and data collection

We examined four risk allocation frameworks—Direct Summation (DS), Probabilistic Union Deduplication (PUD), Risk Population Combination (RPC), and Agent-Based Framework (ABF)—each with both static and dynamic implementations (see Section: Risk allocation frameworks). To demonstrate how these frameworks can be applied in epidemiological modelling, we embedded each within a deterministic compartmental transmission model of TB epidemiology in China and used this model to evaluate the impact of the NTSP from 2025 to 2035.

Population data included dynamic population size, age structure (stratified into three age groups: 0–14, 15–64, and ≥65 years), crude birth rate, and crude death rate from 2010 to 2035, were extracted from the World Bank’s Demographic and Health Surveys database ^22^. Both historical estimates (2010–2023) and future projections (2024–2035) reflect China’s ongoing demographic transition, characterized by accelerated population aging and declining fertility. Age-specific contact patterns were derived from empirical social mixing survey data ^23^. A high-resolution national-level contact matrix (85×85 age groups) was weighted by population age distribution and aggregated into 3×3 matrices corresponding to the defined age strata. This approach allowed the model to accurately capture the age-assortative mixing patterns observed in the Chinese population.

National estimates of TB incidence and mortality were sourced from the World Health Organization (WHO) Global TB Program ^24^. Age-specific notification data (reported cases) from 2010 to 2023 were sourced from the Chinese Center for Disease Control and Prevention (China CDC). Key parameters influencing TB natural history, clinical course of TB disease, and high-risk populations (individuals living with HIV and diabetes mellitus) were integrated through literature reviews, WHO database and China CDC database. Detailed contact matrix processing algorithms, parameter values, and data sources are provided in the Supplementary materials (**Section S1**, **Table S1**–**S2**).

### Risk allocation frameworks

We proposed four analytical frameworks, each comprising both static and dynamic versions, for screening populations with multiple overlapping high-risk factors (**Fig. 1 A**). The core of these frameworks is to estimate the probability that an individual or population group will be covered and detected through screening according to their risk multipliers. Multiple risk factors, including being a close contact of an active TB case, HIV infection, diabetes mellitus, and a prior history of active TB, were considered. Each risk factor is assigned a specific multiplier to adjust its association with both: (i) the TB infection rate multiplier (Multiplier 1) and (ii) the active TB prevalence multiplier among infected individuals (Multiplier 2). Detailed construction procedures of multi-risk frameworks and information about risk multipliers are provided in the Supplementary Materials (**Sections S1**–**S4, and Table S3**–**S4**). Brief descriptions of the four methods are as follows:

i. Direct Summation (DS) Method: The simplest approach, which directly sums the number of individuals requiring screening across each high-risk group. This method does not account for overlapping populations and may therefore result in double counting.
ii. Probabilistic Union Deduplication (PUD) Method: Building upon the DS method, this approach applies the probabilistic union formula ^28^ to correct for duplicate counts arising from overlapping risk groups, thereby yielding a more accurate estimate.
iii. Risk population combination (RPC) Method: This method assumes that the combined effect of multiple risk factors on an individual’s overall risk follows a multiplicative relationship. It simulates possible synergistic effects by constructing all possible combinations of risk factors.
iv. Agent-Based Framework (ABF) Method: The most advanced approach, which simulates the screening process by generating a population of “agents” with individualized risk profiles ^29^. It inherently handles population overlap and possible risk interactions at the micro level, producing estimates that most closely resemble real-world dynamics.

**Figure 1.**
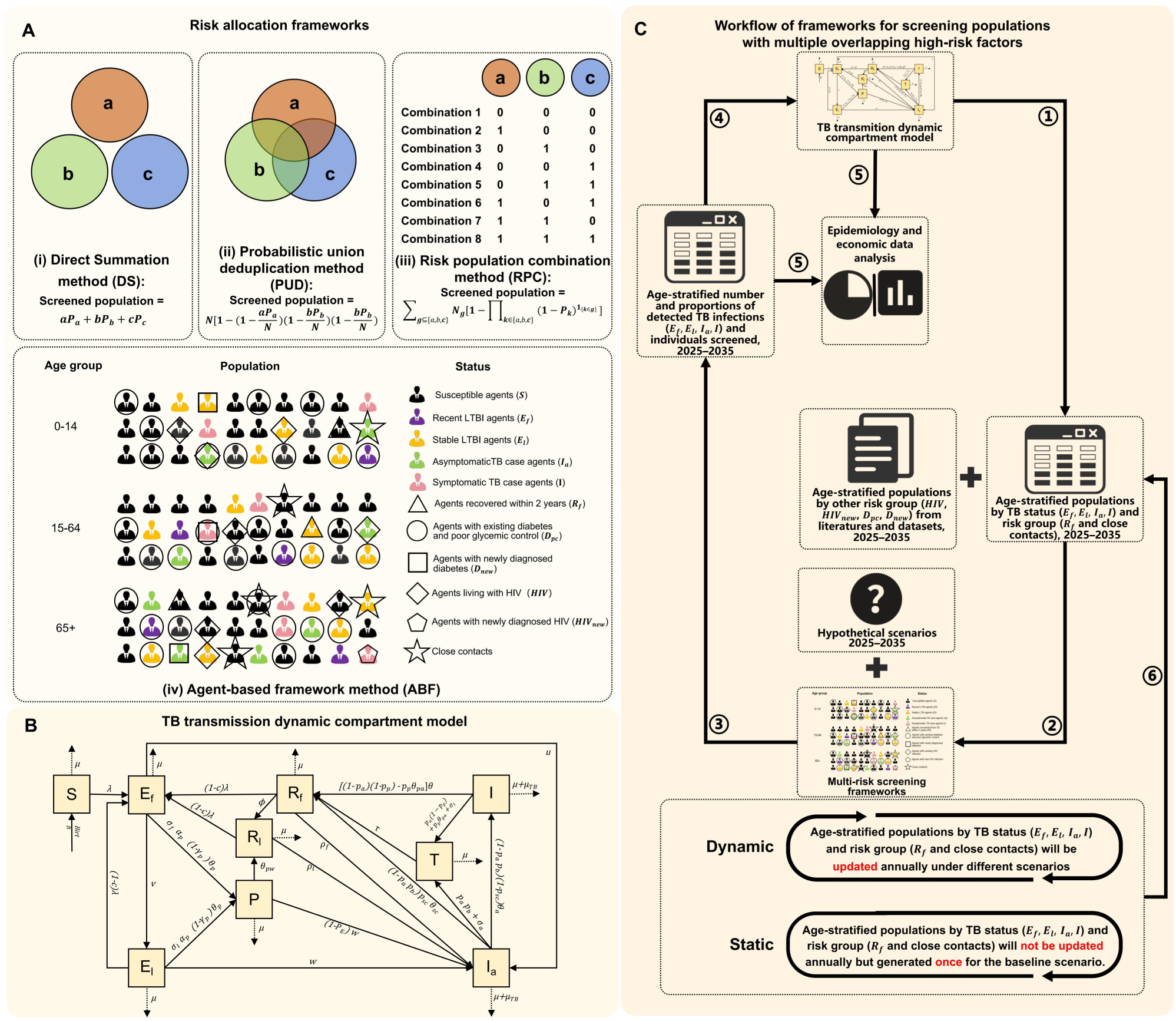
Overview of study framework. **(A)** Risk allocation frameworks. **(B)** Structure of TB transmission dynamic compartment model.**(C)** Workflow of frameworks for screening populations with multiple overlapping high-risk factors. *S:*susceptible population; *E_f_*: recent LTBIs; *E_f_*l: remote LTBIs; *P*: LTBIs protected by preventive therapy; *I_a_*: asymptomatic active TB cases; *I*: symptomatic active TB cases; *T*: TB cases under treatment; *R_f_*: recently recovered individuals; *R_l_*: remotely recovered individuals.

Both the RPC and ABF methods incorporate an interaction parameter γ to capture the combined effects of overlapping risk factors: γ = 1 corresponds to purely multiplicative effects (main analysis), while γ > 1 (synergistic) and γ < 1 (sub-multiplicative) are explored in sensitivity analyses. Crucially, both methods also strictly bound state allocations within the total population pool, preventing estimates from logically exceeding actual case limits.

The flowchart in **Fig. 1 C** outlines this workflow of frameworks for screening populations with multiple overlapping high-risk factors. In total, interventions are simulated at individual or population group level using the aforementioned risk allocation frameworks, with screening effects converted into screening parameters and fed back into the compartmental model.

First, using a TB transmission dynamics compartmental model, we generated age-stratified population sizes categorized by TB status ( *E_f_, E_l_, I, I_a_*) and risk type (*R_f_* and close contacts). Subsequently, additional age-stratified high-risk population data (e.g., individuals living with HIV or diabetes mellitus) are obtained from literature and datasets. These data are integrated into the multi-risk screening framework together with specified intervention target populations and coverage for intervention scenarios from 2025 to 2035. The framework estimates age-stratified number and proportions of detected TB infections (*E_f_, E_l_, I, I_a_*) and individuals screened, and proportions of detected TB infections are then fed as screening parameters into the compartmental model. The entire process operates organically. Finally, epidemiological and economic analyses are conducted on outputs generated by both the compartmental model and the multi-risk screening framework.

The framework supports two operational versions: a non-real-time version and a real-time version. The non-real-time version performs post-simulation calculations using static baseline outputs, thereby failing to capture cumulative intervention effects timely. In contrast, the real-time approach dynamically integrates screening-effect calculations into each iteration of the compartmental model, allowing intervention effects for each year to be recalculated based on the updated population state from the previous year. This closed-loop feedback mechanism enables the model to more accurately reflect the dynamic evolution and long-term impacts of interventions in reducing TB burden, avoiding the systematic overestimation of intervention benefits inherent in the non-real-time version.

### TB transmission dynamic compartment model

We established a TB transmission dynamics model (**Fig. 1 B**) using a compartmental framework similar to previous studies ^25, 26^ and stratified by three age groups (0-14, 15-64, 65+ years old). It includes 9 epidemiological compartments: susceptible population (*S*, recent LTBIs (*E_f_*), remote LTBIs (*E_l_*), LTBIs protected by preventive therapy (*P*), asymptomatic active TB cases (*I_a_*), symptomatic active TB cases (*I*), TB cases under isolation and treatment (*T*), recently recovered individuals (*R_f_*), and remotely recovered individuals (*R_l_*). These compartments are governed by ordinary differential equations (ODEs) that incorporate demographic processes, infection progression, treatment, and relapse. Detailed information about ODEs, calculation of close contacts, and initial values of 9 epidemiological compartments are provided in the Supplementary materials (**Section S2-S3, and Table S5**).

To ensure the model accurately reflects China’s TB epidemiological characteristics, we calibrated key unknown parameters utilized nationwide incidence, mortality and age-specific notifications data from China between 2010 and 2023. The optimization problem was solved using algorithms from the R package “optimx” ^27^, yielding optimal parameter estimates. Detailed technical specifications regarding the model calibration and parameter estimation are provided in the Supplementary materials (**Section S4**).

### Intervention scenarios

Specific interventions include: (i) Screening for LTBIs combined with TB preventive treatment (TPT) targeted at: Close contacts of active TB cases; Individuals newly diagnosed with HIV (*HIV_new_*) Individuals living with HIV (*HIV*); (ii) Active TB case finding and anti-TB treatment (ACF): Close contacts; *HIV_new_*; *HIV*; Recently recovered individuals (*R_f_*); Individuals aged 65 and above; Individuals newly diagnosed with diabetes mellitus (*D_new_*); Individuals living with diabetes mellitus and poor glycemic control (*D_pc_*).

Two scenarios were evaluated: (*i*) The baseline scenario, which maintains the 2024 levels of intervention coverage and extends the observed long-term trends through 2035; and (*ii*) The NTSP scenario, which incorporates all the aforementioned specific interventions and gradually scales up their implementation. Detailed intervention target populations and coverage are summarized in **Table 1**.

**Table 1.**
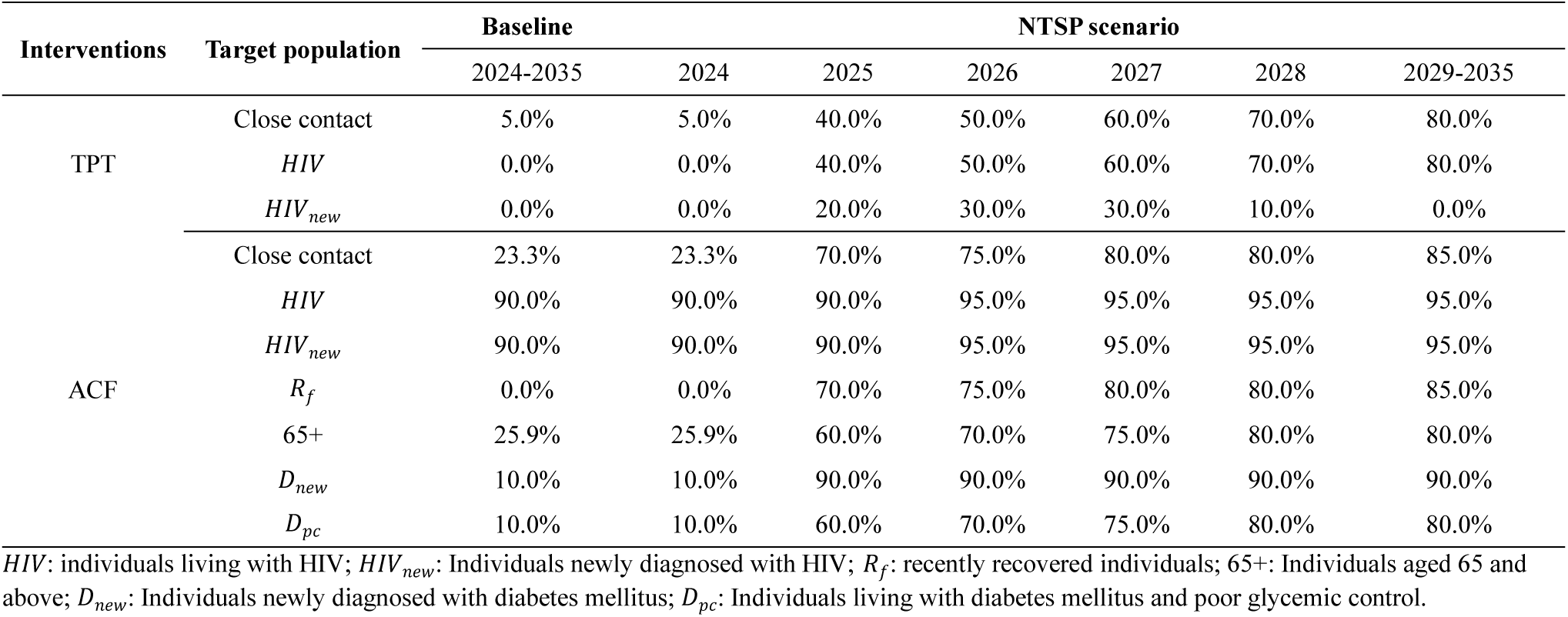
Interventions and coverage for target populations under baseline scenario and the NTSP scenario.

### Data analysis

To illustrate the calibration performance of the constructed TB transmission dynamic compartmental model, we plotted the model fit alongside historical epidemiological data. Because the DS, PUD, and RPC methods do not suffer from stochastic bias, we only validated the robustness and accuracy of the ABF method. We generated 100 agent-based populations representing the year 2025 and calculated the prevalence ratio of TB status between high-risk groups and the general population.

Under the NTSP scenario, we applied different risk allocation frameworks with both static and dynamic versions to the target populations. Comparative results stratified by year and age group were presented using the following indicators: (*i*) Intervention cost indicators: number of TB infection (*E_f_, E_l_, I, I_a_*) detected and number of individuals screened; (*ii*) Screening parameters: proportion of TB infection screened; (*iii*) Intervention benefits indicators: number of TB cases and deaths avoided. The terms “intervention cost” and “intervention benefits” are used as descriptive indicators inspired by health-economic evaluation frameworks ^30, 31^, where the respective quantities could correspond to potential costs or benefits if linked to unit values. In this study, these indicators serve primarily for comparative assessment across scenarios.

To further evaluate the robustness of the results, a sensitivity analysis was performed by varying screening intensity from 20% to 100% of the NTSP target levels. Simulations were conducted under different multi-risk screening frameworks with both static and dynamic versions. For each coverage level, the cumulative numbers of averted TB cases and deaths were recalculated to examine the stability of the model outcomes across a range of implementation intensities. Additionally, a sensitivity analysis on the interaction parameter γ (ranging from 0.25 to 4) was conducted to assess the impact of varying synergistic or sub-multiplicative effects among overlapping risk factors (Supplementary Materials, Fig. S6).

All analyses were conducted using R software version 4.5.2, and the major packages included tidyverse, deSolve, ggplot2, optimx and so on.

## Results

### TB transmission model calibration and ABF method verification

Evaluated across multiple validation dimensions, our transmission dynamic compartment model robustly reproduces national historical trends of TB. The model-estimated national incidence closely aligns with WHO estimates for 2010-2023 (Supplementary Materials, **Fig. S1 A and B**). Importantly, the model accurately captures age-specific notification patterns across fifteen population strata nationwide (Supplementary Materials, **Fig. S1 C**). This comprehensive calibration provides a strong foundation for subsequent assessments of multi-risk screening framework. To further validate the robustness and accuracy of the ABF method, we generated 100 agent-based populations representing the year 2025. Results demonstrated that the prevalence ratios of TB status between high-risk groups and the general population in these simulated agent populations were highly consistent with the predefined values (Supplementary Materials, **Fig. S2**).

### Intervention cost indicators comparative results

Our analysis revealed substantial differences in the intervention cost indicators—specifically, the number of TB infections (*E_f_, E_l_, I, I_a_*) detected and the number of individuals screened—among the four multi-risk screening frameworks with static and dynamic versions under the NTSP scenario. Except for the year 2025, the real-time implementation of all four methods consistently produced lower annual cost metrics than their non-real-time versions.

For the number of *I* and *I_a_* cases detected, a consistent hierarchy was observed across both non-real-time and real-time versions: DS > PUD > RPC ≈ ABF. In contrast, the trend for *E_f_* and *E_l_*infections detected varied by implementation version. Under the static version, all four methods identified a similar number of *E_f_* and *E_f_, E_l_, I, I_a_* infections (DS ≈ PUD ≈ RPC ≈ ABF). Under the real-time version, however, the yields followed an opposite gradient: DS < PUD < RPC ≈ ABF.

For the ACF measure, the number of individuals screened in the static version was highest for the DS method (DS > PUD ≈ RPC ≈ ABF). This pattern was reversed in the dynamic version, where DS required the fewest screenings (DS < PUD < RPC ≈ ABF). A similar reversal was observed for the TPT measure. While the required screening numbers were comparable across methods in the static version (DS ≈ PUD ≈ RPC ≈ ABF), the dynamic version showed the pattern of DS < PUD < RPC ≈ ABF.

By 2035, the cumulative number of *I* cases identified of total population was projected to be 8,752,036 (static) vs. 3,456,205 (dynamic) for DS; 6,566,610 vs. 3,033,926 for PUD; 5,410,843 vs. 2,762,577 for RPC; and 5,412,200 vs. 2,756,400 for ABF. The numbers for *I_a_, E_f_, E_l_*, and the populations screened by ACF and TPT are detailed in **Fig.2 A**–**D.**

**Figure 2.**
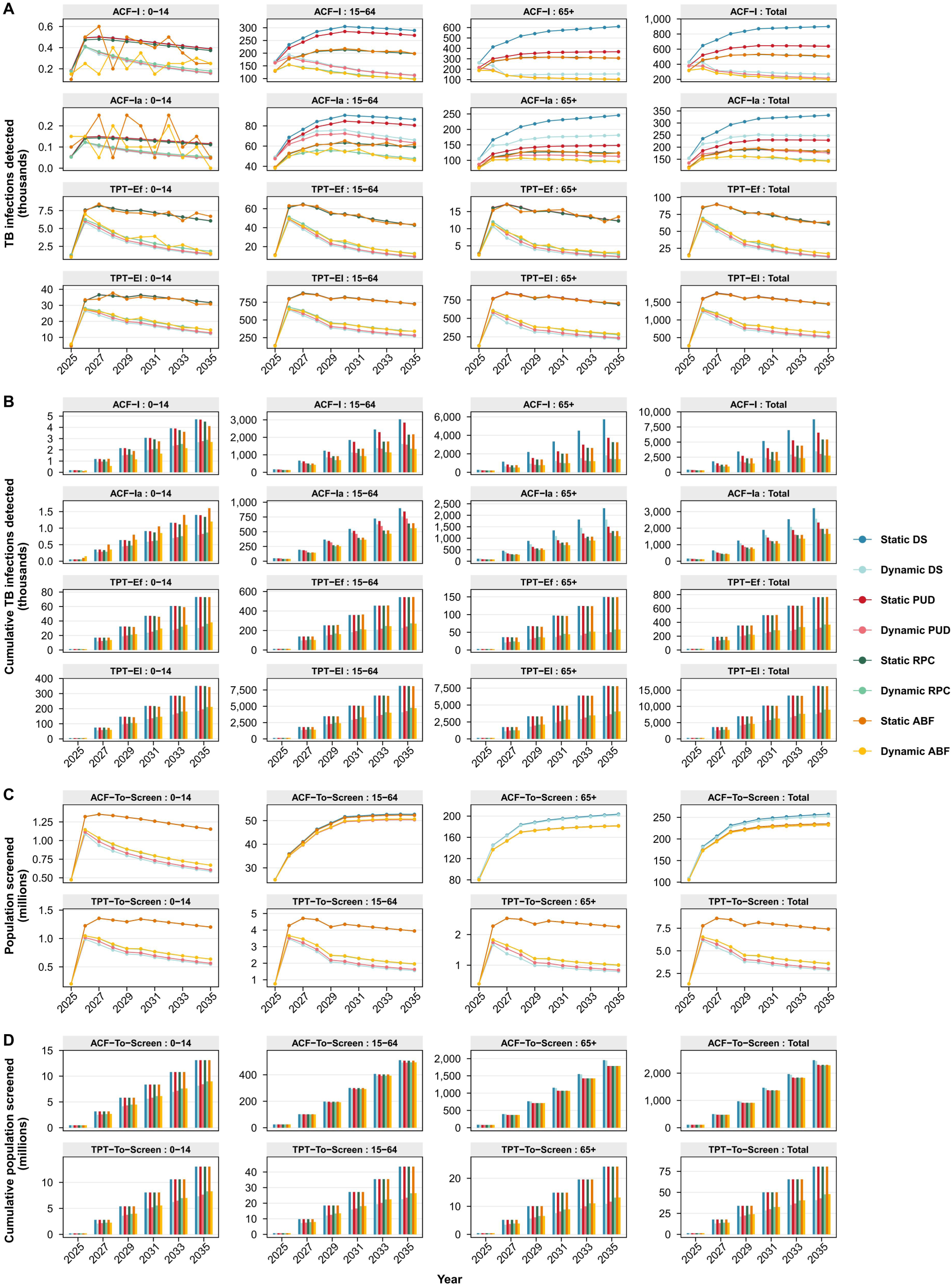
Number of TB infection (*E_f_, E_l_, I, I_a_*) detected and number of individuals screened by age group, 2025-2035. **(A)** Number of TB infection (*E_f_, E_l_, I, I_a_*) detected. **(B)** Cumulative number of TB infection (*E_f_, E_l_, I, I_a_*) detected. **(C)** Number of individuals screened. **(D)** Cumulative number of individuals screened. *I*: symptomatic active TB cases; *I_a_*: asymptomatic active TB cases; *E_f_*: recent LTBIs; *E_l_*: remote LTBIs.

### Screening parameters comparative results

For the proportion of *I* and *I_a_* cases detected, a consistent hierarchy was observed across both non-real-time and real-time versions: DS > PUD > RPC ≈ ABF. Within each multi-risk screening framework, the non-real-time version produced slightly higher values than the real-time version, although the difference was minimal. For the proportion of *E_f_* and *E_l_* infections detected, the four risk allocation frameworks in the static version showed little variation (DS ≈ PUD ≈ RPC ≈ ABF). However, a consistent hierarchy was observed across dynamic versions: DS < PUD < RPC ≈ ABF (Supplementary Materials, **Fig. S3 A and B**).

### Intervention benefits indicators comparative results

Under the baseline scenario without intervention, the projected annual numbers of new TB cases and deaths by 2035 are 612,255 and 17,384, respectively. Under the NTSP scenario, the national TB burden declines markedly, although substantial heterogeneity remains across the four multi-risk screening frameworks, indicating clear differences in Intervention benefits indicators. Specifically, compared with the baseline scenario, the NTSP scenario yields fewer averted TB cases for the dynamic versions than for the static versions of all four risk allocation frameworks, and the differences between risk allocation frameworks also exist (DS > PUD > RPC ≈ ABF). By 2035, the cumulative averted cases in the total population are DS (485,250 vs 359,361), PUD (473,329 vs 357,272), RPC (450,479 vs 349,321), and ABF (450,584 vs 348,868) for real-time versus non-real-time version, respectively. In contrast, in the NTSP scenario, the number of deaths averted relative to the baseline are similar between the dynamic and static versions within each risk allocation frameworks, whereas the differences across risk allocation frameworks are substantial (DS > PUD > RPC ≈ ABF). By 2035, the cumulative averted deaths total 97,664 vs 92,371 vs 82,681 vs 82,884 (real-time) and 95,869 vs 90,784 vs 81,075 vs 80,927 (non-real-time) for DS, PUD, RPC, and ABF, respectively. Detailed data are shown in **Fig.3 A**–**D.**

**Figure 3.**
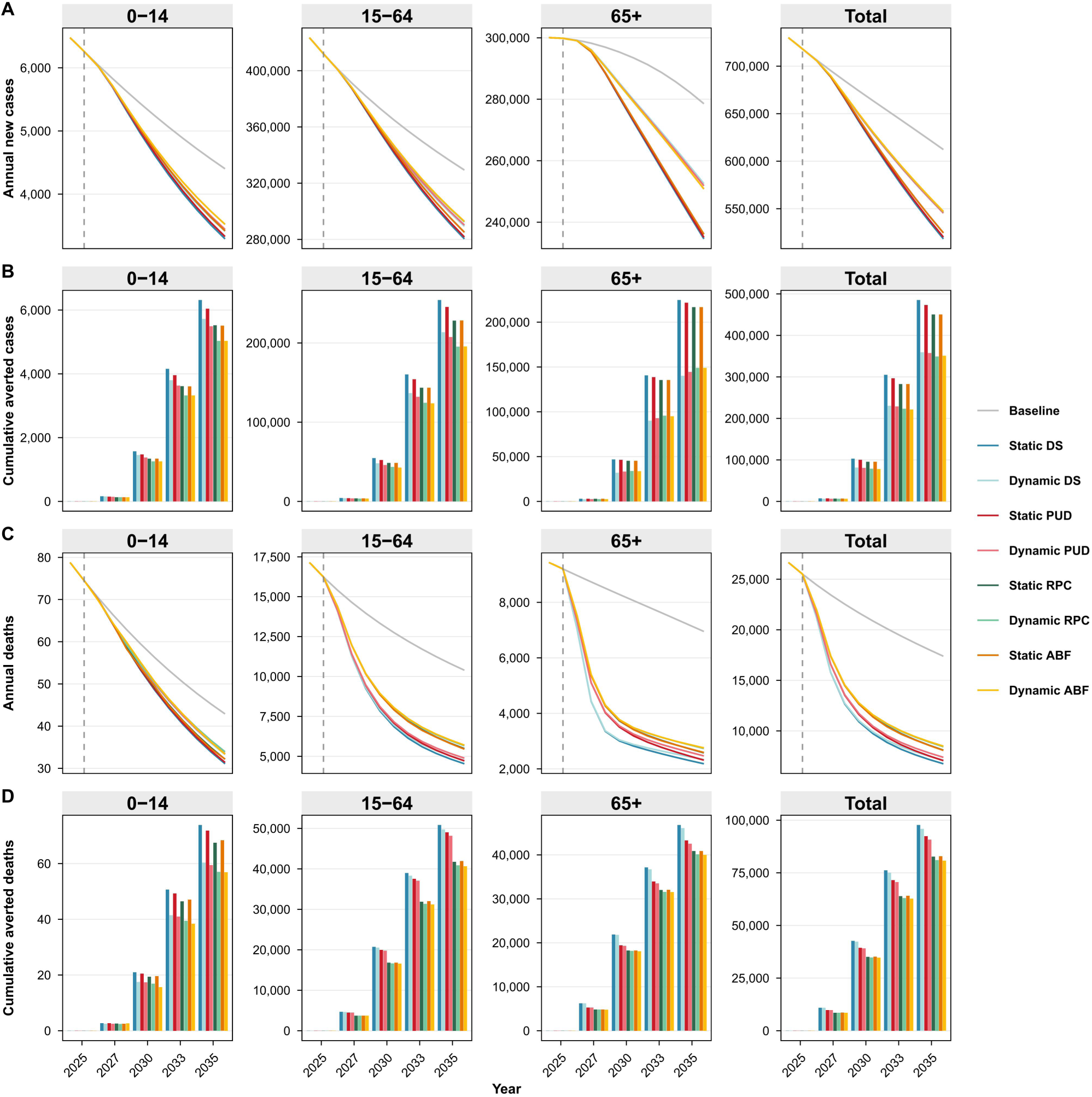
TB incidence and mortality projection under NTSP scenarios of different multi-risk screening methods by age group, 2025-2035. **(A)** Number of TB annual new cases. **(B)** Cumulative number of averted TB cases. **(C)** Number of annual TB deaths. **(D)** Cumulative number of averted TB deaths.

### Sensitivity analysis

Results from the sensitivity analysis indicate that the cumulative numbers of averted TB cases and deaths were robust across all screening intensity levels (20%–100%) of the NTSP scenario (Supplementary Materials, **Fig. S4 A and B**). Across both non-real-time and real-time implementations, these cumulative outcomes consistently followed the hierarchy DS > PUD > RPC ≈ ABF among the four multi-risk screening frameworks. The magnitude of cumulative TB cases averted was more strongly influenced by whether the framework operated in a real-time mode version. In the sensitivity analysis of the interaction parameter γ, the proportion of TB infections detected varied across different γ values (0.25 to 4) across age groups. When γ is large, the overestimation attributable to the methodological crudeness of the DS and PUD methods is correspondingly attenuated. (Supplementary Materials, **Fig. S6**).

## Discussion

Accurate evaluation of screening strategies targeting multiple high-risk populations simultaneously must address the practical challenge of overlapping individual risks. However, most existing models overlook this complexity, focusing instead on single-risk groups or relying exclusively on community-level coverage parameters. Such simplifications detach analyses from real-world policy conditions, potentially lead to biased estimates of program benefits and distorted cost-effectiveness assessments. To provide a structured approach for navigating these challenges, this study presents and compares four risk allocation frameworks that integrate probabilistic and individual-level modeling with traditional transmission-dynamic models. We demonstrate the frameworks’ application through a case study on TB prevention and control within the context of China’s national policy.

### Interpretation of findings

The observed discrepancies across the four risk allocation frameworks under the static version primarily reflect structural methodological differences in how population heterogeneity and risk overlap are characterized. In contrast, the divergence observed in the dynamic version captures the composite effect of these methodological variations interacting with the system’s dynamic feedback loops. Consequently, the variations within a multi-risk screening framework between the two version isolate the specific impact of the dynamic feedback mechanism on intervention outcomes.

Analysis of the static version

In the static version, the number of *I* and *I_a_* cases detected followed a consistent hierarchy: DS > PUD > RPC ≈ ABF. This gradient is attributable to the structural methodologies of each multi-risk screening framework. The DS method produced the highest estimates due to the systematic double-counting of overlapping high-risk individuals. The PUD method yielded lower estimates by employing probabilistic de-duplication, assuming independent acquisition of TB status between risk factors. The RPC and ABF frameworks, furthermore, strictly limits the proportion of state assignments within the total population and the case pool, thereby mathematically avoiding the logical overestimation that occurs in the DS and PUD methods. These methods provided estimates that most closely approximate reality, resulting in a yield slightly lower than the PUD method. Regarding the screened population for ACF, the results (DS > PUD ≈ RPC ≈ ABF) align with theoretical expectations: DS simply sums the target groups, while the other three methods apply probabilistic de-duplication to varying degrees of precision.

For *E_f_* and *E_l_* infections detected and the screened population for TPT, all four risk allocation frameworks produced broadly similar results (DS ≈ PUD ≈ RPC ≈ ABF). This convergence arises from the demographic characteristics of the TPT target population—primarily close contacts and people living with HIV—which constitute a small fraction of the total population (0.7% and 0.1% respectively in 2025). Given the minimal overlap between these specific groups and their relatively low risk of TB infection (Risk Multiplier 1), the methodological differences in overlap correction had a negligible impact on the results.

#### Dynamics in the dynamic version and comparative assessment

Under the dynamic version, where TB status (*E_f_, E_l_, I, I_a_*) and risk group (*R_f_* and close contacts) are updated annually, distinct dynamic trends emerged. Notably, the number of LTBIs detected (*E_f_* and *E_l_*) showed a reversed trend compared to active cases detected (*I* and *I_a_*): DS < PUD < RPC ≈ ABF. This shift is driven by the rapid depletion of the close contact pool—the primary target for TPT. As the intervention proceeds, the “stock” of close contacts is exhausted more efficiently in models with higher granular fidelity, creating differential depletion rates across frameworks.

When comparing the static and dynamic versions, we observed that the number of deaths averted remained remarkably stable between the static and dynamic versions within the same multi-risk screening framework. This stability stems from the composition of the ACF target population, which is dominated by structural risk groups (e.g., the older and individual with diabetes) whose population sizes are relatively independent of TB transmission dynamics. Since screening coverage for these groups remained constant, the proportion of detected active cases (*I* and *I_a_*) remained consistent (Supplementary Materials, **Fig. S3**), ensuring a stable mortality impact.

In sharp contrast, the number of cases averted differed significantly between the two versions. This discrepancy is primarily driven by the dynamic nature of the TPT target population. In the dynamic version, the pool of close contacts shrinks over time due to the intervention and changing epidemic dynamics, leading to a progressive reduction in the detection and treatment of latent infections (*E_f_* and *E_l_*) compared to the static assumptions of the static version (**Fig. 1 A-B**). This underscores the necessity of dynamic feedback modeling when evaluating interventions targeting rapidly depleting subpopulations.

### Characteristics and performance of multi-risk screening framework

Based on methodological principles and findings from China’s TB screening model, we evaluate four frameworks for screening overlapping high-risk populations, summarizing their respective advantages, limitations, and utility. These frameworks present a spectrum of trade-offs between realism and complexity, informing the selection of the optimal method for policy analysis (**Table 2**).

**Table 2.** Comparison of different risk allocation frameworks.

| Characteristics | DS | PUD | RPC | ABF |
| --- | --- | --- | --- | --- |
| Overlap handling | Direct sum of subgroup counts; no explicit overlap handling. | Uses a union-probability formula to avoid double-counting across subgroups. | Explicit risk-profile strata; applies union of screening channels within each risk stratum. | Individual-level representation; each person can be eligible through multiple channels but is counted only once per intervention round. |
| Assumption | Does not explicitly represent the joint distribution of risk factors or their interaction; implicitly accepts double-counting as an approximation. | i. Mutual independence of risk factors at the population level; ii. No interaction between risk factors on infection and disease progression. | i. Mutual independence of risk factors at the population level; ii. Multiplicative interaction of risk factors on infection and disease progression. | Same independence and multiplicative-risk assumptions as the multiplicative method, but implemented at the individual level with finite-population stochastic sampling. |
| State allocation | Macro-level state proportions applied to total counts within each risk group. | Macro-level state proportions applied after overlap adjustment. | States allocated to each risk-profile stratum using risk-based weighting. | States assigned to individuals via weighted sampling based on risk multipliers. |
| Screening probability | Group-specific coverage applied directly to eligible counts. | Union probability across screening groups under independence. | Union probability across screening channels within each risk stratum. | Agent-level stochastic selection according to target coverage; each individual screened at most once per intervention round. |
| Strengths | Conceptually simple; very fast; minimal storage requirements. | Avoids inflation of screened numbers due to overlap; still fast and lightweight. | Captures synergy of multiple risk factors while remaining fast and lightweight; strictly bounds state allocations within the total population pool (prevents logically exceeding actual case limits). | Most realistic: captures both overlap and multi-risk synergy at the individual level; naturally constrained by the simulated population size; incorporates stochastic variability; can serve as a reference or validation standard. |
| Limitations | Overestimates screening volume and yield when subgroups overlap substantially; unbounded by the total case pool, potentially estimating more cases than actually exist. | Does not capture interaction between risk factors; lacks a global constraint for the total case pool, which may lead to overestimation of yields in extreme scenarios. | Cannot represent within-stratum heterogeneity or stochastic uncertainty. | High computation and storage demand; stochastic outputs require multiple runs for stable estimates. |
| Overall preference | Not recommended for main analyses; can be used for quick checks when overlap is minor. | Use with caution as an intermediate benchmark when independence assumptions are plausible. | Strongly recommended for routine analysis. | Recommended as a detailed benchmark and for uncertainty or distributional analyses; may be used selectively due to computational cost. |

DS: Direct Summation; PUD: Probabilistic Union Deduplication; RPC: Risk population combination; ABF: Agent-Based Framework

By simply summing the sizes of individual risk groups, the DS method lacks a global constraint on the total population and case pools. Consequently, it systematically overestimates the target population size, intervention cost indicators, and intervention benefit indicators. Its utility is therefore restricted to preliminary, conservative scenario exploration in contexts where risk group overlap is negligible.

The PUD method offers a significant improvement by employing the probabilistic union formula to correct for duplicate counts arising from overlapping risk groups. This method assumes that risk factors are independent at the population level and no interaction between risk factors on infection and disease progression. It represents a pragmatic balance for policy modeling, being lightweight yet providing a more accurate estimate of the eligible population than the DS method. However, its primary limitations lie in its lack of a global constraint for the total case pool—which may still lead to overestimation of yields in extreme scenarios—and its inability to capture synergistic or sub-multiplicative interactions between risk factors.

The RPC method advances further by explicitly modeling the combined effect of multiple risk factors. By stratifying the population according to risk profiles and applying a multiplicative model to adjust disease progression risks, it not only accounts for overlap but also captures the increased per-capita yield of screening individuals with concurrent risks—a better reflection of biological reality. Crucially, this explicit stratification strictly bounds state allocations within the total population and case pools, mathematically preventing the logical overestimations seen in the DS and PUD methods. Achieving this with only a modest increase in methodological complexity without requiring additional computational resources over the PUD method, RPC serves as our recommended approach for routine analysis.

Finally, the ABF method constitutes the most granular and realistic approach. Operating at the level of individual agents with unique risk profiles, it naturally handles overlap and complex risk interactions, and global population constraints without relying on aggregate-level independence assumptions. Its stochastic nature further enables the examination of outcome distributions and uncertainty ranges. Therefore, the ABF method could serve as a gold standard for validating simpler models. However, as an individual model, it possesses inherent limitations that its high computational cost and the requirement for multiple simulations to ensure stability ^32^, rendering it less practical for rapid policy exploration or extensive sensitivity analyses.

### Methodological advancements over existing screening frameworks

Compared with other modelling studies, this research employs a more refined and efficient approach to addressing multiple high-risk populations.

One common approach is mutual exclusion by definition. For example, a study modeling TPT scale-up in 12 high-burden countries assessed two priority groups: People living with HIV (PLHIV) and Household Contacts (HHCs) ^33^. To avoid double-counting, the authors explicitly defined the HHC cohort as “HIV-negative contacts”, thereby rendering the two groups mutually exclusive. While this allowed for the direct summation of health outcomes from both cohorts, it essentially bypasses the intersection of risks, failing to capture the specific dynamics or synergistic efficiencies of treating individuals who fall into both categories (e.g., HIV-positive contacts).

Another study employing logic analogous to this study’s PUD framework has addressed overlapping high-risk populations, such as the prioritization of attributes for tuberculosis prevention and treatment programs ^34^: This involves separately estimating the population sizes of HIV-infected individuals and household contacts of tuberculosis patients, estimating the overlap proportion using epidemiological methods, and then applying inclusion-exclusion principles to eliminate duplicate individuals, yielding a non-duplicated total population size. This approach effectively avoided double counting of population size and coverage rates but failed to assess interactions between risk factors or consider the potential scenario where dual-risk populations might face higher disease progression risks or derive greater benefits from interventions.

A multinational economic evaluation study on tuberculosis prevention treatment (Brazil, Georgia, Kenya, South Africa) adopted an approach to defining high-risk groups ^35^: each country selected only one primary high-risk group (e.g., prison populations, drug treatment populations, slum and refugee camp populations, and high-prevalence subpopulations) and estimated its population size and tuberculosis prevalence. Within the transmission dynamics equations, the general population and this high-risk group were parameterized and fitted separately. While structurally simple, this approach fails to account for duplicate population size calculations when multiple risks coexist, nor does it capture interactions between risk factors. Consequently, in scenarios involving parallel interventions for multiple high-risk groups, it may underestimate resource wastage or overestimate intervention benefits. Furthermore, compartmental models inherently limit the number of subpopulations that can be modelled: each additional compartment requires distinct epidemiological and intervention parameters, significantly increasing model dimensionality and calibration complexity. In practice, this restricts analysis to a limited number of risk groups with reliable data; increasing stratification often leads to identifiability issues, unstable fits, or reliance on untested assumptions.

In contrast, the proposed multi-risk screening framework of present study, especially strongly recommended RPC method, enables precise estimation of intervention cost indicators and intervention benefits indicators during simultaneous screening of multiple high-risk groups, with the consideration of interaction of multi-risks and avoiding an explosion in parameter space compared to multi-subpopulation compartment model. Consequently, it maintains computational feasibility and robustness even in highly stratified policy scenarios.

### Strengths and limitations

To our knowledge, this study represents the first systematic evaluation of screening frameworks specifically designed for populations with multiple overlapping risk factors. We have introduced four risk allocation frameworks—ranging from simple summation to agent-based simulations—and synthesized their theoretical characteristics and implementation pathways. This work offers a scalable reference framework for future health economic evaluations involving heterogeneous risk profiles.

Despite these contributions, our study has several limitations. First, all four frameworks assume that risk factors are independently distributed across the population. In reality, risk factors can cluster—for example, individuals in overcrowded housing may also face greater exposure to air pollution—and the joint frequency of overlapping risk factors is rarely known with precision. Our frameworks do not resolve this underlying data gap; rather, they provide a structured way to allocate risks under the assumption of independence, pending the availability of richer empirical data on risk factor co-occurrence. Second, our frameworks assume that the combined effect of multiple risk factors on TB disease risk follows a multiplicative relationship. While this assumption aligns with current biological evidence for key TB risk drivers (e.g., HIV and diabetes), the true nature of risk interactions—whether multiplicative, additive, or otherwise—remains uncertain in the absence of direct empirical evidence on the joint effects of overlapping risks. Third, the RPC and ABF frameworks incorporate an interaction parameter γ to capture departure from multiplicativity, but we have not systematically validated the hybrid approach against a transmission model that is fully stratified by multiple and overlapping risk factors with explicit clustering effects. Such cross-validation, while computationally demanding, would strengthen confidence in the framework’s accuracy and is an important direction for future work. Fourth, this study was conducted using China as a case study for TB; while the core methodological insights are not constrained by geography or disease, the generalizability of specific quantitative findings to other settings requires further investigation.

## Conclusion

This study presents and systematically compares four potential screening frameworks for populations with multiple overlapping risk factors. Our findings demonstrate that traditional modeling approaches, which overlook individual risk overlap and dynamic feedback mechanisms, tend to over- or underestimate intervention effectiveness and resource requirements. Through comparative methodological analysis, we find that the RPC method offers the most favorable balance between estimation accuracy and computational tractability among the frameworks evaluated, and we propose the dynamic RPC method as the preferred tool for routine analysis where multimorbidity and intersectional risks exist. Furthermore, our findings highlight the pivotal role of dynamic feedback loops: ignoring the dynamic depletion of high-risk pools can lead to substantial errors in projecting long-term incidence reductions. Although focusing on TB control in China, the modular design of the proposed frameworks allows for adaptation to other disease settings characterized by multimorbidity and intersectional risks. Importantly, we acknowledge that all four frameworks share common limitations, including the assumptions of independently distributed risk factors and multiplicative combined effects, which future work with richer data on overlapping risks could address. Taken together, this work provides a transparent methodological comparison to inform the optimization of public health screening strategies and resource allocation across complex risk landscapes.

## Supporting information

supplementary_Risk_allocation_Framework

## Data Availability

All data produced in the present study are available upon reasonable request to the authors

## Abbreviations

ABF: Agent-Based Framework
ACF: Active Case Finding
CDC: Center for Disease Control and Prevention
DS: Direct Summation
HHCs: Household Contacts
HIV: Human Immunodeficiency Virus
LTBI: Latent Tuberculosis Infection
MLE: Maximum Likelihood Estimation
RPC: Risk population combination
NTSP: National Tuberculosis Strategic Plan (2025–2035)
ODEs: Ordinary Differential Equations
PLHIV: People Living with HIV
PUD: Probabilistic Union Deduplication
TB: Tuberculosis
TPT: TB Preventive Treatment
WHO: World Health Organization

## Acknowledgments

We express our sincere gratitude to all participants.

## Declarations

### Authors’ contributions

**Wenyong Zhou:** Conceptualization, Data Curation, Software, Formal analysis, Writing - Original Draft. **Zexuan Wen:** Conceptualization, Data Curation, Software, Formal analysis, Writing - Original Draft. **Tao Li**: Data Curation, **Xiaoqiu Liu**: Data Curation, **Canyou Zhang**: Data Curation, **Yunzhou Ruan**: Data Curation, **Hui Zhang**: Data Curation. **Nimalan Arinaminpathy:** Conceptualization, Resources, Methodology, Writing - Review & Editing, Project administration, Funding acquisition. **Weibing Wang:** Conceptualization, Resources, Methodology, Writing - Review & Editing, Project administration, Funding acquisition. All authors read and approved the final manuscript.

### Data and code availability

The data and R code are available upon request from the authors.

### Ethics approval and consent to participate

Not applicable.

### Consent for publication

All authors have approved the manuscript for submission.

