## supplementary_Risk_allocation_Framework for "Comparing screening frameworks for populations with multiple overlapping high-risk factors: A case of tuberculosis screening in China"

* Corresponding author:

### Content

**Section S1 Contact Patterns** ... 3

**Section S2 Direct Summation method** ... 4

**Section S3 Probabilistic union deduplication method** ... 6

**Section S4 Risk population combination method** ... 8

**Section S5 Agent-based multi-risk screening framework** ... 11

**Section S6: Transition dynamics governed by ordinary differential equations** ... 15

**Section S7 Calculation of close contacts** ... 17

**Section S8 TB transmission dynamic compartment model calibration and parameter estimation** ... 18

**Figure S1. TB transmission dynamic compartment model calibration to WHO estimated incidence(A), mortality (B) and age-stratified notification data(C), 2010-2023.** ... 19

**Figure S2. Prevalence ratio of tuberculosis status between high-risk groups and the general population of 100 generated population.** ... 20

**Figure S3. Proportion of TB infection ($Ef, El, I, Ia$) detected by age group, 2025-2035.** ... 21

**Figure S4. Cumulative numbers of averted TB cases and deaths across all screening intensity levels (20%–100%) of the NTSP scenario by age group, 2025-2035.** ... 22

**Figure S5. Population dynamics of nine compartments.** ... 23

**Figure S6. Proportion of TB infection ($Ef, El, I, Ia$) detected by age group under different** $\gamma$ (0.25 to 4), **2025-2035.** ... 26

**Table S1 TB transmission dynamic compartment model parameters and descriptions.** ... 27

**Table S2 Description and sources of data.** ... 29

**Table S3 TB infection rate multiplier (Multiplier 1).** ... 30

**Table S4 Active TB prevalence multiplier among infected individuals (Multiplier 2).** ... 31

**Table S5 Initial values of 9 epidemiological compartments by age group of 2010.** ... 32

**References:** ... 33

#### Section S1 Contact Patterns

We define:

$G_i$, $G_j$: Set of ages in row group i (participants) and column group j (contacts)

$m_{xy}$: Original contact rate between age x and age y (85×85)

$P_x$, $P_y$ : Population count at age x and age y

$C_{ij}$: Aggregated weighted contact rate for group i contacting group j

$$C_{ij} = \frac{\sum_{x \in G_i} \left( \frac{\sum_{y \in G_j} m_{xy} P_y}{\sum_{y \in G_j} P_y} \right) P_x}{\sum_{x \in G_i} P_x}$$

Final age-specific contact matrices:

$$CM = \begin{bmatrix} 0.4951 & 0.1330 & 0.0292 \\ 0.0906 & 0.2301 & 0.0405 \\ 0.0598 & 0.1193 & 0.0781 \end{bmatrix}$$

where matrix elements represent contact rates between age groups 0-14, 15-64, and 65+ years.

#### Section S2 Direct Summation method

##### Notation and definitions

We define:

$j$: index for risk groups.

$y$ : index for year, $y \in \{2025, \ldots, 2035\}$.

$a$: index for age group, $a \in \{0-14, 15-64, 65+\}$.

$k$: total number of risk groups for ACF.

$k_{TPT}$: total number of risk groups for TPT.

$n_j$: the number of people in risk group $j$.

$\theta_j$: coverage proportion for the respective risk group for ACF.

$\theta_j^{\mathrm{TPT}}$: coverage proportion for the respective risk group for TPT.

$I$: symptomatic active TB cases.

$I_a$: asymptomatic active TB cases.

$E_f$: recent LTBIs.

$E_l$: remote LTBIs.

$p_I$: proportion of $I$ in each stratum $(y, a)$.

$p_{I_a}$: proportion of $I_a$ in each stratum $(y, a)$.

$p_{E_f}$: proportion of $E_f$ in each stratum $(y, a)$.

$p_{E_l}$: proportion of $E_l$ in each stratum $(y, a)$.

$r_{1,j}$: TB infection rate multiplier for risk factor $j$ (Multiplier 1).

$r_{2,j}$: Active TB prevalence multiplier among infected individuals for risk factor $j$ (Multiplier 2).

##### Calculation of the intervention cost indicators

Number of individuals screened via ACF screening of each stratum $(y, a)$:

$$N_{\mathrm{ACF}} = \sum_j^k n_j \theta_j$$

Number of individuals screened via TPT screening of each stratum $(y, a)$:

$$N_{\mathrm{TPT}} = \sum_j^k n_j \theta_j^{\mathrm{TPT}}$$

Number of $I$ detected via ACF screening of each stratum $(y, a)$:

$$I_{\mathrm{ACF}} = \sum_{j}^{k} n_j \theta_j p_I r_{1,j} r_{2,j}$$

Number of $I_a$ detected via ACF screening of each stratum $(y, a)$:

$$I_{a,\mathrm{ACF}} = \sum_{j}^{k} n_j \theta_j p_{I_a} r_{1,j} r_{2,j}$$

Number of $E_f$ detected via TPT screening of each stratum $(y, a)$:

$$E_{f,\mathrm{TPT}} = \sum_{j}^{k_{\mathrm{TPT}}} n_j \theta_j^{\mathrm{TPT}} p_{E_f} r_{1,j}$$

Number of $E_l$ detected via TPT screening of each stratum $(y, a)$:

$$E_{l,TPT} = \sum_{j}^{k_{TPT}} n_j \theta_j^{TPT} p_{E_l} r_{1,j}$$

#### Section S3 Probabilistic union deduplication method

**Notation and definitions**

We define:

$j$: index for risk groups

$y$ : index for year, $y \in \{2025, \ldots, 2035\}$.

$a$: index for age group, $a \in \{0-14, 15-64, 65+\}$.

$T$: total population in each stratum $(y, a)$.

$k$: total number of risk groups for ACF.

$k_{TPT}$: total number of risk groups for TPT.

$n_j$: the number of people in risk group $j$.

$\theta_j$: coverage proportion for the respective risk group for ACF.

$\theta_j^{\text{TPT}}$: coverage proportion for the respective risk group for TPT.

$I$: symptomatic active TB cases.

$I_a$: asymptomatic active TB cases.

$E_f$: recent LTBIs.

$E_l$: remote LTBIs.

$p_I$: proportion of $I$ in each stratum $(y, a)$.

$p_{I_a}$: proportion of $I_a$ in each stratum $(y, a)$.

$p_{E_f}$: proportion of $E_f$ in each stratum $(y, a)$.

$p_{E_l}$: proportion of $E_l$ in each stratum $(y, a)$.

$I_{\text{total}}$: total numbers of $I$ in each stratum $(y, a)$.

$I_{a,\text{total}}$: total numbers of $I_a$ in each stratum $(y, a)$.

$E_{f,\text{total}}$: total numbers of $E_f$ in each stratum $(y, a)$.

$E_{l,\text{total}}$: total numbers of $E_l$ in each stratum $(y, a)$.

$r_{1,j}$: TB infection rate multiplier for risk factor $j$ (Multiplier 1).

$r_{2,j}$: Active TB prevalence multiplier among infected individuals for risk factor $j$ (Multiplier 2).

**Calculation of the intervention cost indicators**

Number of individuals screened via ACF of each stratum $(y, a)$:

$$N_{ACF} = T\left[1 - \prod_{j=1}^{k}\left(1 - \frac{n_j\theta_j}{T}\right)\right].$$

Number of individuals screened via TPT of each stratum $(y, a)$:

$$N_{TPT} = T\left[1 - \prod_{j=1}^{k_{TPT}}\left(1 - \frac{n_j\theta_j^{TPT}}{T}\right)\right].$$

Proportions of $I$ detected via ACF screening of each stratum $(y, a)$:

$$p_j^{(I)} = \frac{n_j\theta_j p_I r_{1,j} r_{2,j}}{I_{total}},$$

$$I_{ACF} = I_{total}\left[1 - \prod_{j=1}^{k}\left(1 - p_j^{(I)}\right)\right].$$

Proportions of $I_a$ detected via ACF screening of each stratum $(y, a)$:

$$p_j^{(I_a)} = \frac{n_j\theta_j p_{I_a} r_{1,j} r_{2,j}}{I_{a,total}},$$

$$I_{a,ACF} = I_{a,total}\left[1 - \prod_{j=1}^{k}\left(1 - p_j^{(I_a)}\right)\right].$$

Number of $E_f$ detected via TPT screening of each stratum $(y, a)$:

$$p_j^{(E_f)} = \frac{n_j\theta_j^{TPT} p_{E_f} r_{1,j}}{E_{f,total}},$$

$$E_{f,TPT} = E_{f,total}\left[1 - \prod_{j=1}^{k_{TPT}}\left(1 - p_j^{(E_f)}\right)\right].$$

Number of $E_l$ detected via TPT screening of each stratum $(y, a)$:

$$p_j^{(E_l)} = \frac{n_j\theta_j^{TPT} p_{E_l} r_{1,j}}{E_{l,total}},$$

$$E_{l,TPT} = E_{l,total}\left[1 - \prod_{j=1}^{k_{TPT}}\left(1 - p_j^{(E_l)}\right)\right].$$

#### Section S4 Risk population combination method

**Notation and definitions**

We define:

$j$: index for risk groups

$y$ : index for year, $y \in \{2025, \ldots, 2035\}$.

$a$: index for age group, $a \in \{0-14, 15-64, 65+\}$.

$T$: total population in each stratum $(y, a)$.

$k$: total number of risk groups for ACF.

$k_{TPT}$: total number of risk groups for TPT.

$n_j$: the number of people in risk group $j$.

$\theta_j$: coverage proportion for the respective risk group for ACF.

$\theta_j^{\mathrm{TPT}}$: coverage proportion for the respective risk group for TPT.

$I$: symptomatic active TB cases.

$I_a$: asymptomatic active TB cases.

$E_f$: recent LTBIs.

$E_l$: remote LTBIs.

$w_i^{(1)}$: TB infection rate multiplier for risk factor $j$ (Multiplier 1).

$w_i^{(2)}$: Active TB prevalence multiplier among infected individuals for risk factor $j$ (Multiplier 2).

$\gamma$: The interaction parameter capturing the combined effects of overlapping risk factors. It is set to $\gamma$=1.0 (purely multiplicative/independent effect) in the main analysis, and varied ($\gamma$>1 for synergistic effects, and $\gamma$<1 for sub-multiplicative effects) in the sensitivity analysis.

**Binary risk factors and their prevalence**

We consider $m$ binary risk factors $R_j$ for TB $(j = 1, \ldots, k)$ , each taking values $R_j \in \{0,1\}$ :

$R_j = 1$ means that the individual has risk factor $j$;

$R_j = 0$ means that the individual does not have this risk factor.

Let $p_j$ denote the proportion of the population with $R_j = 1$ (the marginal prevalence of risk factor $j$ ):

$$p_j = Pr(R_j = 1), j = 1, \ldots, k.$$

**Risk combination generation**

A specific combination of risk factors is represented by the vector $\mathbf{R} = (r_1, ..., r_k)$, where each $r_j \in \{0,1\}$ indicates whether risk factor $j$ is present in that combination. For example, with 3 factors (contact, HIV, diabetes), $\mathbf{R} = (1,0,1)$ could mean "contact = yes, HIV = no, diabetes = yes".

Under the assumption that the $k$ risk factors are independent in the population, the proportion of the population that has exactly the combination **R** is

$$\pi(\boldsymbol{R}) = Pr(R_1 = r_1, ..., R_k = r_k) = \prod_{j=1}^{k} \left[r_j p_j + (1 - r_j)(1 - p_j)\right],$$

except that HIV is split into mutually exclusive "new" and "old" components. Let the HIV-related indicators be $HIV_{new}, HIV_{old} \in \{0,1\}$ with marginal prevalences $p_{HIV_{new}}$ and $p_{HIV_{old}}$, and define

$$p_{HIV_{none}} = 1 - p_{HIV_{new}} - p_{HIV_{old}}.$$

Then in the product above, the single HIV term is replaced by

$$\pi_{HIV}(HIV_{new}, HIV_{old}) = \begin{cases} p_{HIV_{new}}, & (HIV_{new}, HIV_{old}) = (1,0), \\ p_{HIV_{old}}, & (HIV_{new}, HIV_{old}) = (0,1), \\ p_{HIV_{none}}, & (HIV_{new}, HIV_{old}) = (0,0), \\ 0, & (HIV_{new}, HIV_{old}) = (1,1). \end{cases}$$

All other risk factors remain independent and enter $\pi(\mathbf{R})$ as before.

**TB states allocation to each combination**

Relative risks:

$$R_1(\boldsymbol{R}) = \gamma^{(\sum r_j - 1)^+} \prod_{j=1}^{k} \left[r_j w_j^{(1)} + (1 - r_j)\right],$$

$$R_2(\boldsymbol{R}) = \gamma^{(\sum r_j - 1)^+} \prod_{j=1}^{k} \left[r_j w_j^{(2)} + (1 - r_j)\right],$$

with the understanding that both $HIV_{new}$ and $HIV_{old}$ use the same HIV multipliers $w_{HIV}^{(1)}$ and $w_{HIV}^{(2)}$ , but cannot be 1 simultaneously. The positive-part exponent $\left(\sum r_j - 1\right)^+ = \max(0, \sum r_j - 1)$ ensures the interaction term equals 1 for combinations with 0 or 1 risk factor.

$$E_f(\boldsymbol{R}) = E_{f,total} \frac{\pi(\boldsymbol{R}) R_1(\boldsymbol{R})}{\sum_{\boldsymbol{R}} \pi(\boldsymbol{R}) R_1(\boldsymbol{R})},$$

$$E_l(\boldsymbol{R}) = E_{l,total} \frac{\pi(\boldsymbol{R}) R_1(\boldsymbol{R})}{\sum_{\boldsymbol{R}} \pi(\boldsymbol{R}) R_1(\boldsymbol{R})},$$

$$I(\boldsymbol{R}) = I_{total} \frac{\pi(\boldsymbol{R}) R_1(\boldsymbol{R}) R_2(\boldsymbol{R})}{\sum_{\boldsymbol{R}} \pi(\boldsymbol{R}) R_1(\boldsymbol{R}) R_2(\boldsymbol{R})},$$

$$I_a(\boldsymbol{R}) = I_{a,total} \frac{\pi(\boldsymbol{R}) R_1(\boldsymbol{R}) R_2(\boldsymbol{R})}{\sum_{\boldsymbol{R}} \pi(\boldsymbol{R}) R_1(\boldsymbol{R}) R_2(\boldsymbol{R})}.$$

where $HIV_{new}$ and $HIV_{old}$ can have distinct coverages (e.g., $\theta_{HIV_{new}}$ and $\theta_{HIV_{old}}$ ), but remain mutually exclusive in $\pi_{HIV}$ .

**Calculation of the intervention cost indicators**

Screening probabilities:

$$q_{ACF}(\boldsymbol{R}) = 1 - \prod_{j=1}^{k} [1 - r_j\theta_j] ,$$

$$q_{TPT}(\boldsymbol{R}) = 1 - \prod_{j=1}^{k_{TPT}} [1 - r_j\theta_j^{TPT}] ,$$

Number of individuals screened via ACF of each stratum $(y, a)$:

$$N_{ACF} = \sum_{\boldsymbol{R}} \pi(\boldsymbol{R}) T q_{ACF}(\boldsymbol{R}).$$

Number of individuals screened via TPT of each stratum $(y, a)$:

$$N_{TPT} = \sum_{\boldsymbol{R}} \pi(\boldsymbol{R}) T q_{TPT}(\boldsymbol{R}) .$$

Number of $I$ detected via ACF screening of each stratum $(y, a)$:

$$I_{ACF} = \sum_{\boldsymbol{R}} I(\boldsymbol{R}) q_{ACF}(\boldsymbol{R}) .$$

Number of $I_a$ detected via ACF screening of each stratum $(y, a)$:

$$I_{a,ACF} = \sum_{\boldsymbol{R}} I_a(\boldsymbol{R}) q_{ACF}(\boldsymbol{R}) .$$

Number of $E_f$ detected via TPT screening of each stratum $(y, a)$:

$$E_{f,TPT} = \sum_{\boldsymbol{R}} E_f(\boldsymbol{R}) T q_{TPT}(\boldsymbol{R}) .$$

Number of $E_l$ detected via TPT screening of each stratum $(y, a)$:

$$E_{l,TPT} = \sum_{\boldsymbol{R}} E_l(\boldsymbol{R}) T q_{TPT}(\boldsymbol{R}) .$$

#### Section S5 Agent-based multi-risk screening framework

**Notation and Definitions**

We define:

$i$: index for agent

$j$: index for risk groups

$y$ : index for year, $y \in \{2025, ..., 2035\}$.

$a$: index for age group, $a \in \{0-14, 15-64, 65+\}$.

$T$: total population in each stratum $(y, a)$.

$k$: total number of risk groups for ACF.

$k_{TPT}$: total number of risk groups for TPT.

$\theta_j$: coverage proportion for the respective risk group for ACF.

$\theta_j^{\mathrm{TPT}}$: coverage proportion for the respective risk group for TPT.

$I$: symptomatic active TB cases.

$I_a$: asymptomatic active TB cases.

$E_f$: recent LTBIs.

$E_l$: remote LTBIs.

$C$: identified TB contacts

$HIV_{\mathrm{new}}$: newly diagnosed HIV patients

$HIV$: existing HIV patients (total HIV minus new HIV)

$R_f$: Recent recovered individuals

$D_{new}$: newly diagnosed diabetes cases

$D_{pc}$: diabetic patients with poor glycemic control

$w_i^{(1)}$: TB infection rate multiplier for agent $i$.

$w_i^{(2)}$: Active TB prevalence multiplier among infected individuals for agent $i$.

$\gamma$: The interaction parameter capturing the combined effects of overlapping risk factors. It is set to $\gamma$=1.0 (purely multiplicative/independent effect) in the main analysis, and varied ($\gamma$>1 for synergistic effects, and $\gamma$<1 for sub-multiplicative effects) in the sensitivity analysis.

**Initial state allocation**

Population size per stratum (year $y$ , age group $a \in \{0-14, 15-64, 65+\}$):

$$N_{y,a} = \left\lfloor \frac{N\text{-}origin_{y,a}}{10} \right\rfloor, \quad I_{y,a} = \left\lfloor \frac{I\text{-}origin_{y,a}}{10} \right\rfloor, I_{a,y,a} = \left\lfloor \frac{Ia\text{-}origin_{y,a}}{10} \right\rfloor,$$

$$E_{f,y,a} = \left\lfloor \frac{Ef\text{-}origin_{y,a}}{10} \right\rfloor, E_{l,y,a} = \left\lfloor \frac{El\text{-}origin_{y,a}}{10} \right\rfloor, R_{f,y,a} = \left\lfloor \frac{Rf\text{-}origin_{y,a}}{10} \right\rfloor,$$

$$HIV_{y,a} = \left\lfloor \frac{HIV\text{-}origin_{y,a}}{10} \right\rfloor, HIV_{new,y,a} = \left\lfloor \frac{HIVnew\text{-}origin_{y,a}}{10} \right\rfloor, D_{new,y,a} = \left\lfloor \frac{Dinc\text{-}origin_{y,a}}{10} \right\rfloor,$$

$$D_{pc,y,a} = \left\lfloor \frac{Dpc\text{-}origin_{y,a}}{10} \right\rfloor, C_{y,a} = \left\lfloor \frac{C\text{-}origin_{y,a}}{10} \right\rfloor.$$

Agents within each stratum $(y, a)$ are assigned a binary status vector representing TB states, risk factors, and contact status. Each agent $i \in \{1, \dots, N_{y,a}\}$ has status vector:

$$\mathbf{S}_i = \left(I_i, I_{a,i}, E_{f,i}, E_{l,i}, R_{f,i}, HIV_i, HIV_{new,i}, D_{new,i}, D_{pc,i}, C_i\right) \in \{0,1\}^{10}.$$

Constraints:

TB states are mutually exclusive:

$$I_i + I_{a,i} + E_{f,i} + E_{l,i} \leq 1.$$

Newly diagnosed HIV is a subset of HIV:

$$HIV_{new,i} \leq HIV_i.$$

**TB status assignment**

Risk multipliers were used for adjusting weighted sampling for TB status assignment.

Risk multiplier 1 ($w_i^{(1)}$) :

$$w_i^{(1)} = \gamma^{(n_i-1)^+} \times \begin{cases} 0.8 & if\ HIV_i = 1 \\ 1 & otherwise \end{cases} \times \begin{cases} 1.47 & if\ (D_{new_i} = 1) \vee (D_{pc_i} = 1) \\ 1 & otherwise \end{cases} \times \begin{cases} 1.38 & if\ C_i = 1 \wedge a = 0\text{-}14 \\ 1.33 & if\ C_i = 1 \wedge a = 15\text{-}64 \\ 1.71 & if\ C_i = 1 \wedge a = 65+ \\ 1.0 & otherwise. \end{cases}$$

Risk multiplier 2 ($w_i^{(2)}$) :

$$w_i^{(2)} = \gamma^{(n_i-1)^+} \times \begin{cases} 30.2 & if R_{f,i} = 1 \\ 1 & otherwise \end{cases} \times \begin{cases} 15.5 & if HIV_i = 1 \\ 1 & otherwise \end{cases} \times \begin{cases} 7.18 & if (D_{new_i} = 1) \vee (D_{pc_i} = 1) \\ 1 & otherwise. \end{cases} \times \begin{cases} 4.97 & if\ C_i = 1 \\ 1 & otherwise \end{cases}$$

where $n_i = \sum_j I(R_{j,i} = 1)$ is the number of risk factors present for agent $i$. The positive-part exponent $(n_i - 1)^+ = \max(0, n_i - 1)$ ensures the interaction term equals 1 for agents with 0 or 1 risk factor.

For $X \in \{R_f, HIV, D_{new}, D_{pc}, C\}$ , we assign exactly $X_{y,a}$ agents to have $X_i = 1$ (sampling without replacement), consistent with the code:

$$\sum_{i=1}^{N_{y,a}} \mathbb{I}\,(X_i = 1) = X_{y,a}.$$

Among HIV-positive agents:

$$\sum_{i:HIV_i=1} \mathbb{I}(HIV_{new,i} = 1) = min\{HIV_{new,y,a}, HIV_{y,a}\}$$

Define aggregated targets:

$$I_{total,y,a} = I_{y,a} + I_{a,y,a},$$

$$E_{total,y,a} = E_{f,y,a} + E_{l,y,a}.$$

Assign $I_{\text{total}}$ among all agents using weights $w_i^{(1)} w_i^{(2)}$ :

$$\mathbb{P}\left(I_{total,i} = 1\right) \propto w_i^{(1)} w_i^{(2)}, \quad \sum_{i=1}^{N_{y,a}} I_{total,i} = I_{total,y,a}.$$

Assign $E_{\text{total}}$ among agents with $I_{\text{total},i} = 0$ using weights $w_i^{(1)}$ :

$$\mathbb{P}\left(E_{total,i} = 1 | I_{total,i} = 0\right) \propto w_i^{(1)}, \quad \sum_{i=1}^{N_{y,a}} E_{total,i} = E_{total,y,a}.$$

**Status Decomposition**

Decompose $E_{\text{total}}$ into $E_f$ and $E_l$ :

$$\mathbb{P}\left(E_{f,i} = 1 | E_{total,i} = 1\right) = \frac{E_{f,y,a}}{E_{f,y,a} + E_{l,y,a}},$$

$$E_{l,i} = \mathbb{I}(E_{total,i} = 1 \wedge E_{f,i} = 0).$$

Decompose $I_{\text{total}}$ into $I$ and $I_a$ :

$$\mathbb{P}\left(I_i = 1 | I_{total,i} = 1\right) = \frac{I_{y,a}}{I_{y,a} + I_{a,y,a}},$$

$$I_{a,i} = \mathbb{I}\left(I_{total,i} = 1 \wedge I_i = 0\right).$$

**Calculation of the intervention cost indicators**

Let $\mathcal{P}_{y,a}$ be the agent set in year $y$ and age group $a$ . For any indicator $Z_i \in \{0,1\}$ , define the eligible set

$$\Omega_{y,a}(Z) = \left\{i \in \mathcal{P}_{y,a} : Z_i = 1\right\}.$$

For ACF screening, within each age group $a$ , we sample without replacement

$$\widetilde{\Omega}_{y,a,j}^{ACF} \subseteq \Omega_{y,a}(j), \quad \left|\widetilde{\Omega}_{y,a,j}^{ACF}\right| = \theta_j(y) |\Omega_{y,a}(j)|,$$

where $\Omega_{y,a}(Age65)$ is defined as $\mathcal{P}_{y,a}$ when $a = 65 +$ and empty otherwise.

The final ACF screened set is the union across all groups and age groups with deduplication:

$$\Omega_y^{ACF} = \bigcup_a \bigcup_j \widetilde{\Omega}_{y,a,j}^{ACF}.$$

Number of individuals screened via ACF of each stratum $(y, a)$:

$$N_{ACF} = 10\left|\Omega_y^{ACF}\right|.$$

Number of $I$ detected via ACF screening of each stratum $(y, a)$:

$$I_{\mathrm{ACF}} = 10 \sum_{i \in \Omega_y^{ACF} \cap \mathcal{P}_{y,a}} \mathbb{I}(I_i = 1),$$

Number of $I_a$ detected via ACF screening of each stratum $(y, a)$:

$$I_{a,\mathrm{ACF}} = 10 \sum_{i \in \Omega_y^{ACF} \cap \mathcal{P}_{y,a}} \mathbb{I}\left(I_{a,i} = 1\right).$$

For TPT screening, within each age group $a$ , we sample without replacement

$$\widetilde{\Omega}_{y,a,\mathrm{j}}^{TPT} \subseteq \Omega_{y,a}(\mathrm{j}), \quad \left|\widetilde{\Omega}_{y,a,\mathrm{j}}^{TPT}\right| = \theta_{\mathrm{j}}^{TPT}(y)|\Omega_{y,a}(\mathrm{j})|.$$

Deduplicated TPT screened set:

$$\Omega_y^{TPT} = \bigcup_a \bigcup_{\mathrm{j}} \widetilde{\Omega}_{y,a,\mathrm{j}}^{TPT}.$$

Number of individuals screened via TPT of each stratum $(y, a)$:

$$N_{\mathrm{TPT}} = 10\left|\Omega_y^{TPT}\right|.$$

Number of $E_f$ detected via TPT screening of each stratum $(y, a)$:

$$E_{f,\mathrm{TPT}} = 10 \sum_{i \in \Omega_y^{TPT} \cap \mathcal{P}_{y,a}} \mathbb{I}\left(E_{f,i} = 1\right).$$

Number of $E_l$ detected via TPT screening of each stratum $(y, a)$:

$$E_{l,\mathrm{TPT}} = 10 \sum_{i \in \Omega_y^{TPT} \cap \mathcal{P}_{y,a}} \mathbb{I}\left(E_{l,i} = 1\right).$$

#### Section S6: Transition dynamics governed by ordinary differential equations

**Transition dynamics compartments:**

Susceptible population:

$$\frac{dS_i}{dt} = birth + AgeIn_{S_i} - \lambda S_i - \mu S_i - AgeOut_{S_i}$$

Recent LTBIs (within 2 years):

$$\frac{dE_{f_i}}{dt} = AgeIn_{E_{f_i}} + \lambda S_i + (1-c)\lambda\left(E_{l_i} + R_{f_i} + R_{l_i}\right) - \left[u + v + \sigma_f \alpha_p (1-\gamma_p)\theta_p + \mu\right]E_{f_i} - AgeOut_{E_{f_i}}$$

Remote LTBIs (more than 2 years):

$$\frac{dE_{l_i}}{dt} = AgeIn_{E_{l_i}} + vE_{f_i} - \left[w + (1-c)\lambda + \sigma_l \alpha_p (1-\gamma_p)\theta_p + \mu\right]E_{l_i} - AgeOut_{E_{l_i}}$$

LTBIs protected by preventive therapy:

$$\frac{dP_i}{dt} = AgeIn_{P_i} + \left(\sigma_f E_{f_i} + \sigma_l E_{l_i}\right)\alpha_p (1-\gamma_p)\theta_p - \left[(1-P_E)w + \theta_{pw} + \mu\right]P_i - AgeOut_{P_i}$$

Asymptomatic TB cases:

$$\frac{dI_{a_i}}{dt} = AgeIn_{I_{a_i}} + uE_{f_i} + wE_{l_i} + (1-P_E)wP_i + \rho_f R_{f_i} + \rho_l R_{l_i} \\ - [p_a p_b + \sigma_a + (1-p_a p_b)(1-p_{sc})\theta_a + (1-p_a p_b)p_{sc}\theta_{sc} + \mu + \mu_{TB}]I_{a_i} - AgeOut_{I_{a_i}}$$

Symptomatic TB cases:

$$\frac{dI_i}{dt} = AgeIn_{I_i} + (1-p_a p_b)(1-p_{sc})\theta_a I_{a_i} - \left[p_a(1-p_p) + p_p\theta_{pa} + \sigma_I + [(1-p_a)(1-p_p) - p_p\theta_{pa}]\theta + \mu + \mu_{TB}\right]I_i \\ - AgeOut_{I_i}$$

TB cases on treatment:

$$\frac{dT_i}{dt} = AgeIn_{T_i} + (p_a p_b + \sigma_a)I_{a_i} + \left[p_a(1-p_p) + p_p\theta_{pa} + \sigma_I\right]I_i - (\tau + \mu)T_i - AgeOut_{T_i}$$

Recently recovered individuals from active TB cases (within 2 years):

$$\frac{dR_{f_i}}{dt} = AgeIn_{R_{f_i}} + \tau T_i + (1-p_a p_b)p_{sc}\theta_{sc}I_a + [(1-p_a)(1-p_p) - p_p\theta_{pa}]\theta I_i - [\rho_f + \phi + (1-c)\lambda + \mu]R_{f_i} \\ - AgeOut_{R_{f_i}}$$

Remotely recovered individuals from active TB cases (more than 2 years):

$$\frac{dR_{l_i}}{dt} = AgeIn_{R_{l_i}} + \phi R_{f_i} + \theta_{pw}P_i - [\rho_l + (1-c)\lambda + \mu]R_{l_i} - AgeOut_{R_{l_i}}$$

Force-of-infection:

$$\lambda_i(t) = \beta_i \sum_{j=1}^{3} CM_{ij} \frac{I_j(t) + b \cdot I_{aj}(t)}{N_j(t)}$$

where $N_j$ is the population size of population aged 0-14, 15-64, and 65+, $CM_{ij}$ is the age-specific contact rate matrix, $\beta$ is the transmission coefficient, and $b$ scales transmissibility for asymptomatic cases. As a previous study suggests [11], aging and demographic dynamics are modeled with time-dependent birth and mortality rates. The compartmental model employs the following assumptions: (*i*) LTBI populations are categorized based on the timing of initial infection into recent and stable progression phases. (*ii*) LTBIs protected by preventive therapy ($P$) exhibit better immune protection; after waning, they are assumed to have lower relapse risk. (*iii*) Recently recovered individuals ($R_f$) face higher relapse risks compared to remotely recovered individuals ($R_l$). (*iv*) Remote LTBIs ($E_l$), recently recovered individuals ($R_f$), and remotely recovered individuals ($R_l$) have lower reinfection risks. (*v*) All TB cases undergo an asymptomatic TB stage, and asymptomatic TB cases exhibit a lower infectiousness compared to active TB cases.

**Tracking compartments:**

In addition to the core states, the model includes 3 tracking compartments to monitor epidemiological outcomes, such as incidences, notifications, and deaths. These do not affect the main dynamics but allow for calibration against observed data and projection of intervention impacts. The equations for these tracking variables are as:

New TB cases:

$$\frac{dCases_i}{dt} = uE_{f_i} + wE_{l_i} + (1 - P_E)wP_i + \rho_f R_{f_i} + \rho_l R_{l_i}$$

New TB notifications:

$$\frac{dNotifications_i}{dt} = (p_a p_b + \sigma_a)I_{a_i} + \left[p_a\left(1 - p_p\right) + p_p\theta_{pa} + \sigma_I\right]I_i$$

New TB deaths:

$$\frac{dDeaths_i}{dt} = \mu_{TB}(I_{a_i} + I_i)$$

#### Section S7 Calculation of close contacts

$$\begin{bmatrix} C_{0-14} \\ C_{15-64} \\ C_{65+} \end{bmatrix} = \begin{bmatrix} 40 & 0.96 & 1 \\ 2 & 5.52 & 2 \\ 2 & 2 & 3 \end{bmatrix} \times \begin{bmatrix} I_{0-14} \\ I_{15-64} \\ I_{65+} \end{bmatrix}$$

Where $C_i$ represent the total number of close contacts calculated for each age group $i$ , and $I_i$ representing the number of infected individuals in each age group.

#### Section S8 TB transmission dynamic compartment model calibration and parameter estimation

Five key epidemiological parameters were selected for estimation: transmission rate ($\beta$), latent infection progression rate ($u$), latent infection stability rate ($v$), endogenous recurrence rate ($w$), and TB fatality rate ($TB_{\mu}$). To account for age heterogeneity, each parameter was adjusted by age-specific correction factors, resulting in 15 parameters to be estimated.

We estimated parameters using the Maximum Likelihood Estimation (MLE) method [2]. The objective function was constructed as the sum of the log-likelihoods for incidence ($\mathcal{L}_{inc}$), age-specific notifications ($\mathcal{L}_{not}$), and mortality ($\mathcal{L}_{mort}$), assuming normally distributed observation errors:

$$log\mathcal{L}_{total}(\Theta) = log\mathcal{L}_{inc} + log\mathcal{L}_{not} + log\mathcal{L}_{mort}$$

The specific components were defined as follows:

$$\log\mathcal{L}_{inc} = \sum_{t=2010}^{2023} \log\phi\left(D_{inc,t} | M_{inc,t}(\Theta), \sigma_{inc}\right)$$

$$\log\mathcal{L}_{not} = \sum_{t=2010}^{2023}\sum_{a=1}^{3} w_{a,t} \cdot \log\phi\left(D_{not,a,t} | M_{not,a,t}(\Theta), \sigma_{not}\right)$$

Here, $w_{a,t}$ represents the normalized inverse weight derived from the observed proportion of notifications in age group $a$ at year $t$.

$$\log\mathcal{L}_{mort} = W_{mort} \sum_{t=2010}^{2023} \log\phi\left(D_{mort,t} | M_{mort,t}(\Theta), \sigma_{mort}\right)$$

In these equations, $\phi(\cdot)$ denotes the probability density function of the normal distribution, $D$ represents observed data, $M(\Theta)$ represents model output, and $\sigma$ represents the assumed standard deviation. Given the importance of capturing mortality trends for policy assessment, we assigned an additional penalty weight ($W_{mort} = 20$) to the mortality component.

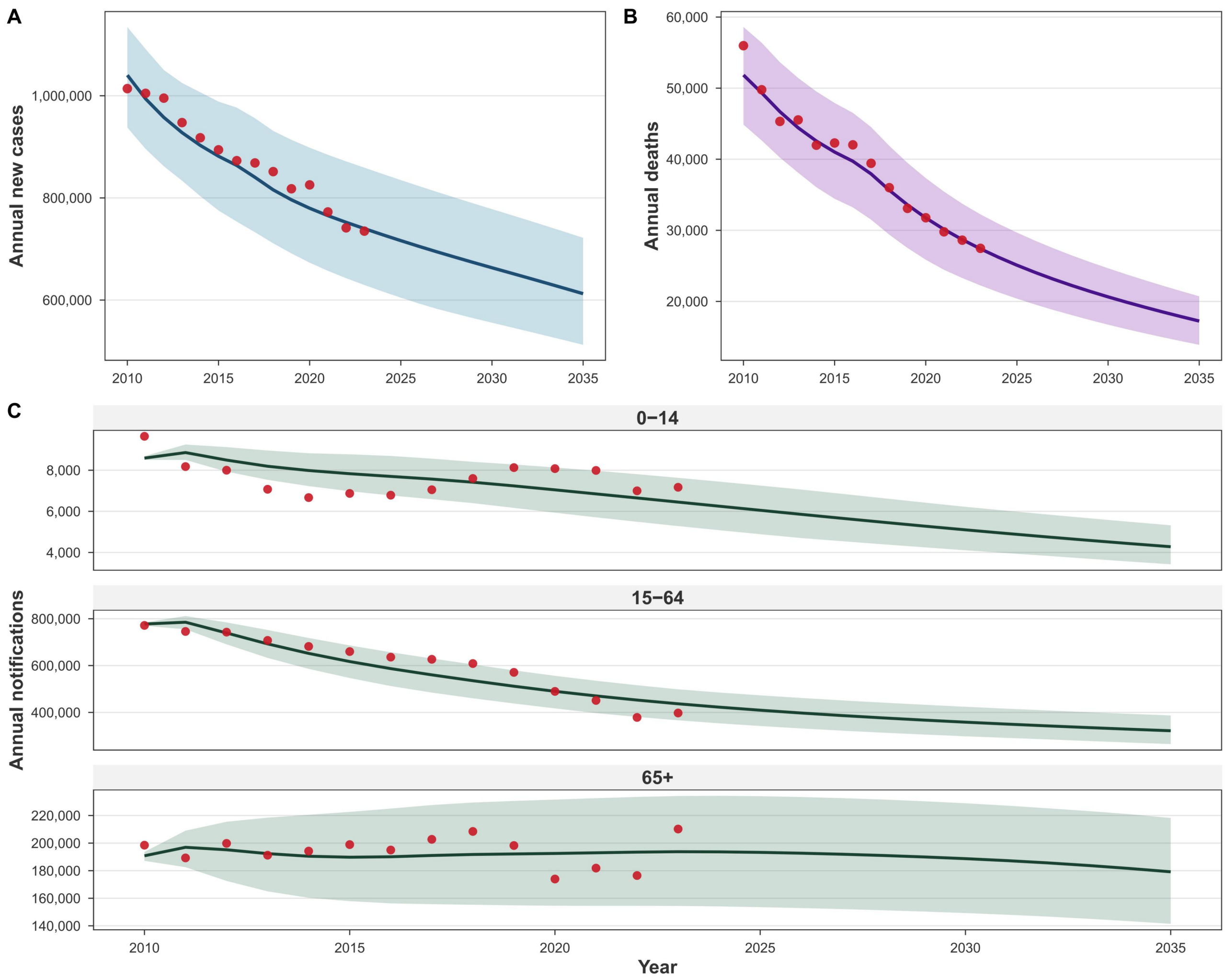


**Figure S1. TB transmission dynamic compartment model calibration to WHO estimated incidence(A), mortality (B) and age-stratified notification data(C), 2010-2023.**

The curve represents the fitted values, while the red dots denote the observed values.

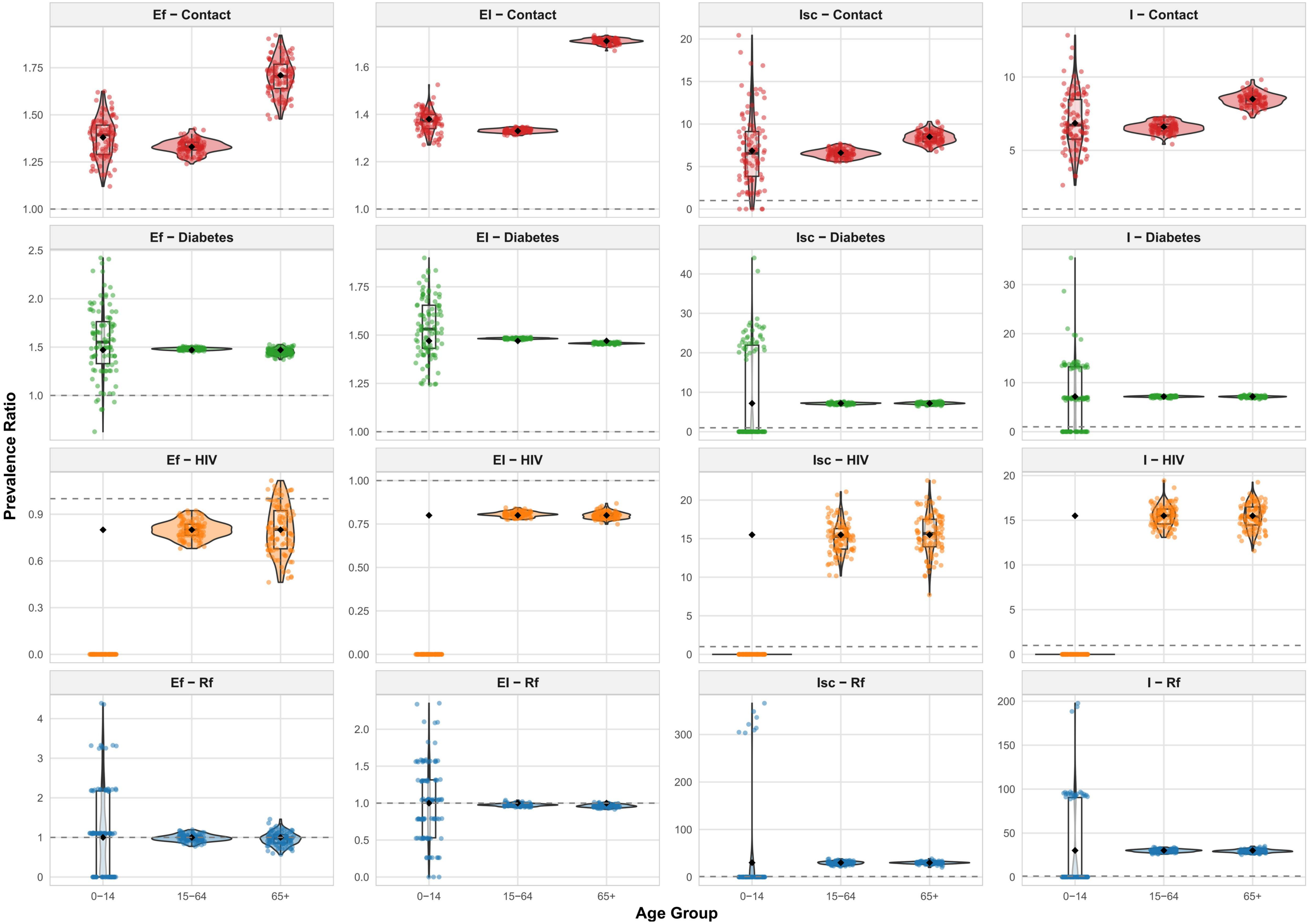


**Figure S2. Prevalence ratio of tuberculosis status between high-risk groups and the general population of 100 generated population.** Black dots denote set values of prevalence ratio, and dots of other colors denote prevalence ratio of generated population.

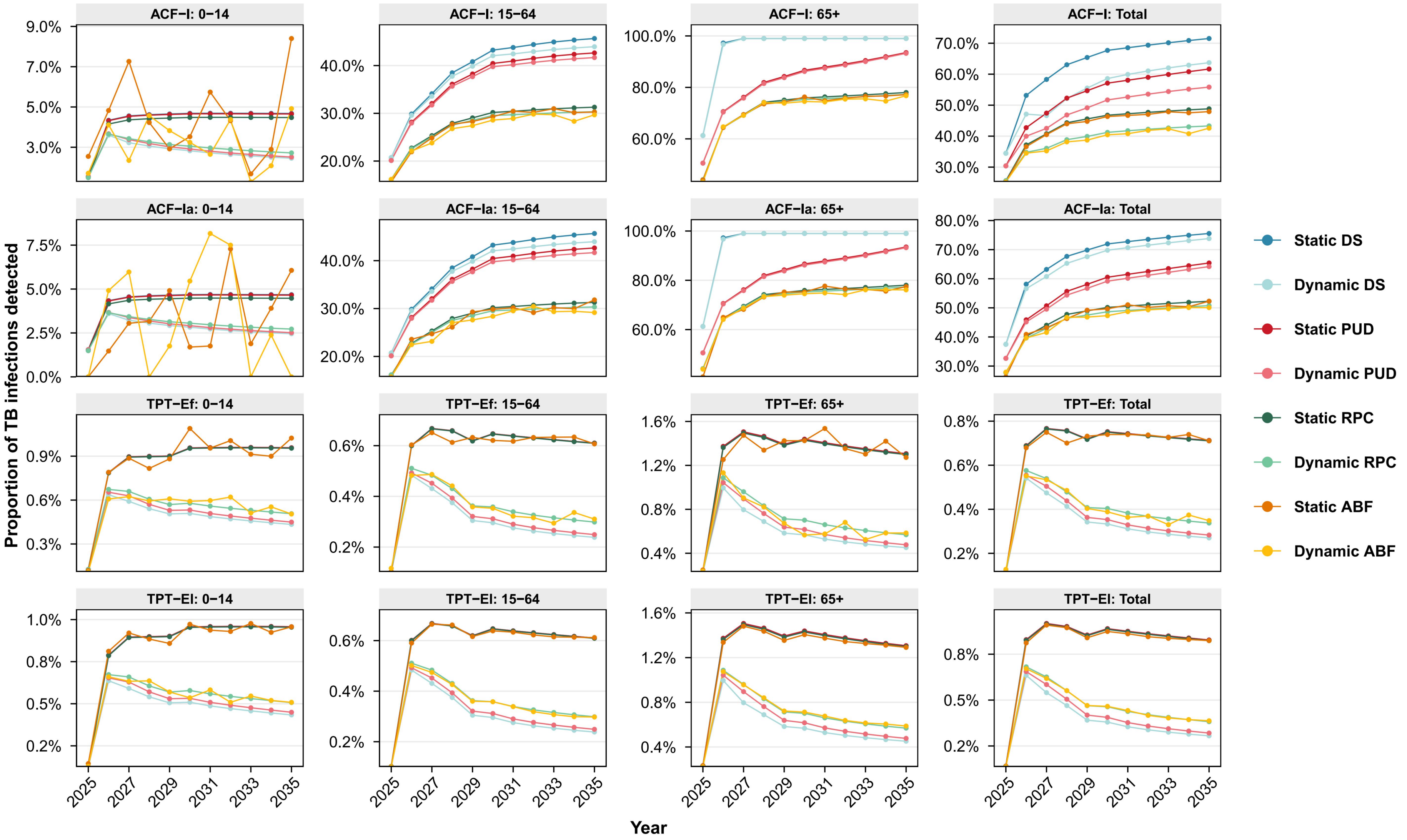


**Figure S3. Proportion of TB infection ($E_f$, $E_l$, $I$, $I_a$) detected by age group, 2025-2035.**

$I_a$: asymptomatic active TB cases; $E_f$: recent LTBIs; $E_l$: remote LTBIs

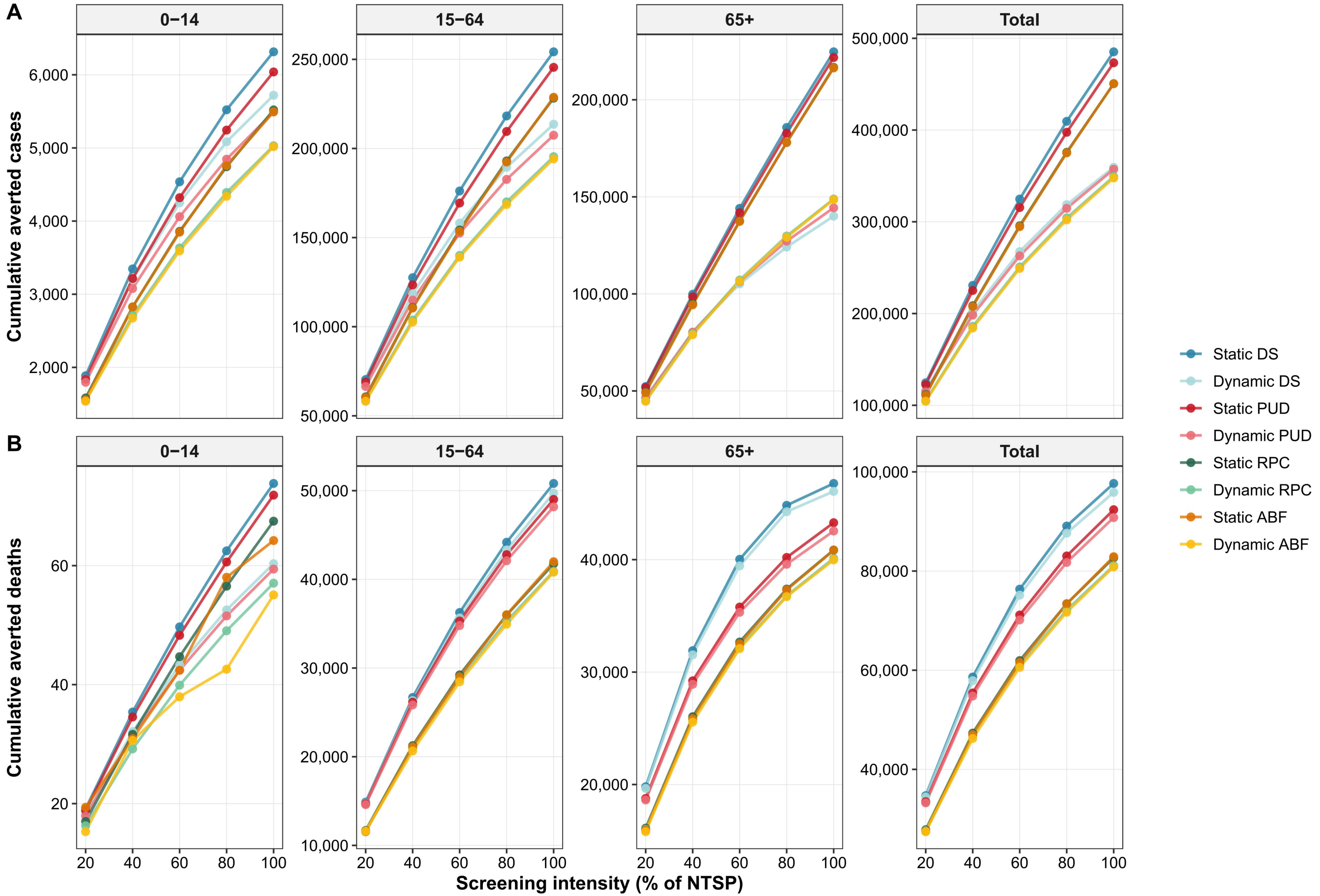


306
307 **Figure S4. Cumulative numbers of averted TB cases and deaths across all screening intensity levels (20%–100%) of the NTSP scenario by age group, 2025-2035.**

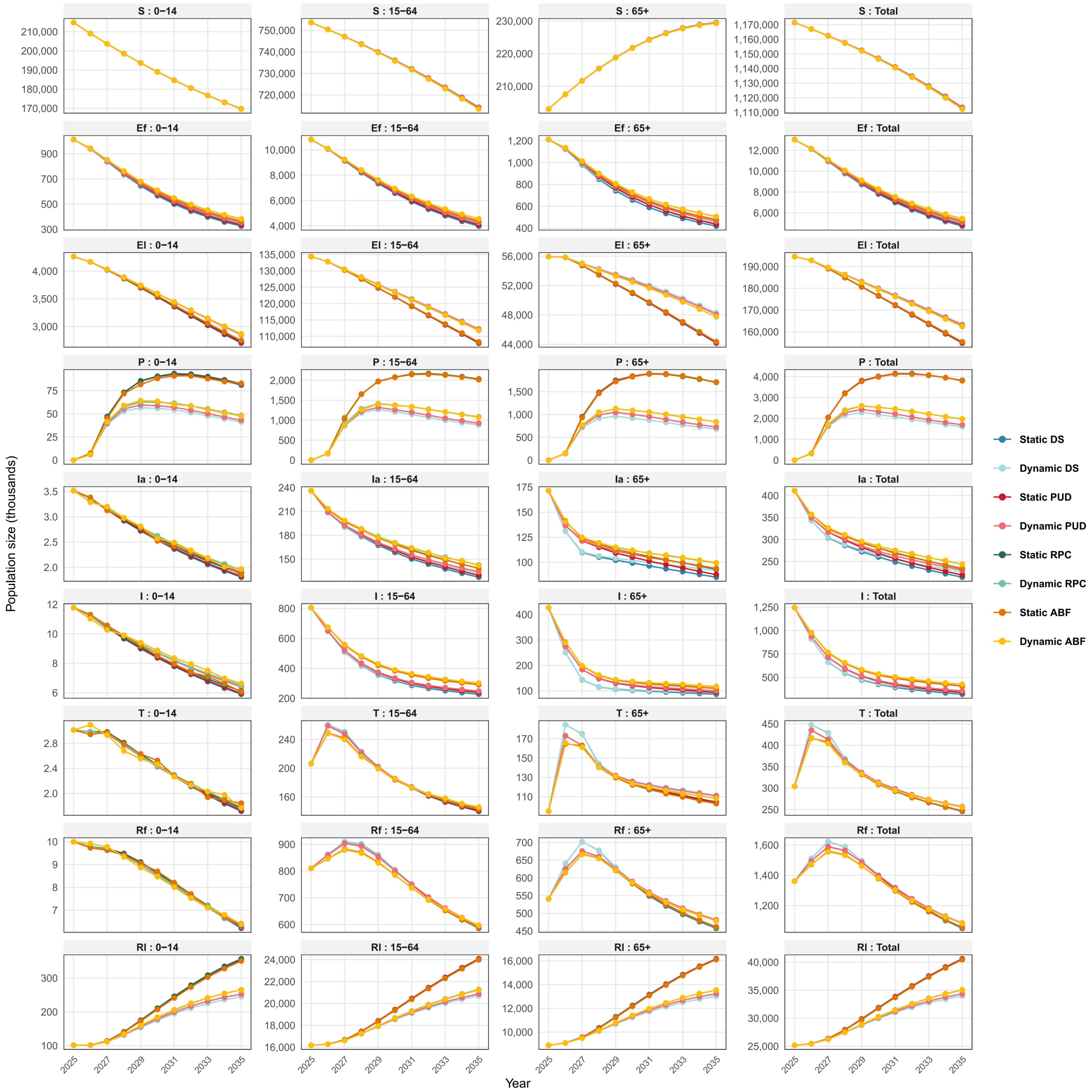


**Figure S5. Population dynamics of nine compartments.**

$S$: susceptible population; $E_f$: recent LTBIs; $E_l$: remote LTBIs; $P$: LTBIs protected by preventive therapy; $I_a$: asymptomatic active TB cases; $I$: symptomatic active TB cases; $T$: TB cases under treatment; $R_f$: recently recovered individuals; $R_l$: remotely recovered individuals.

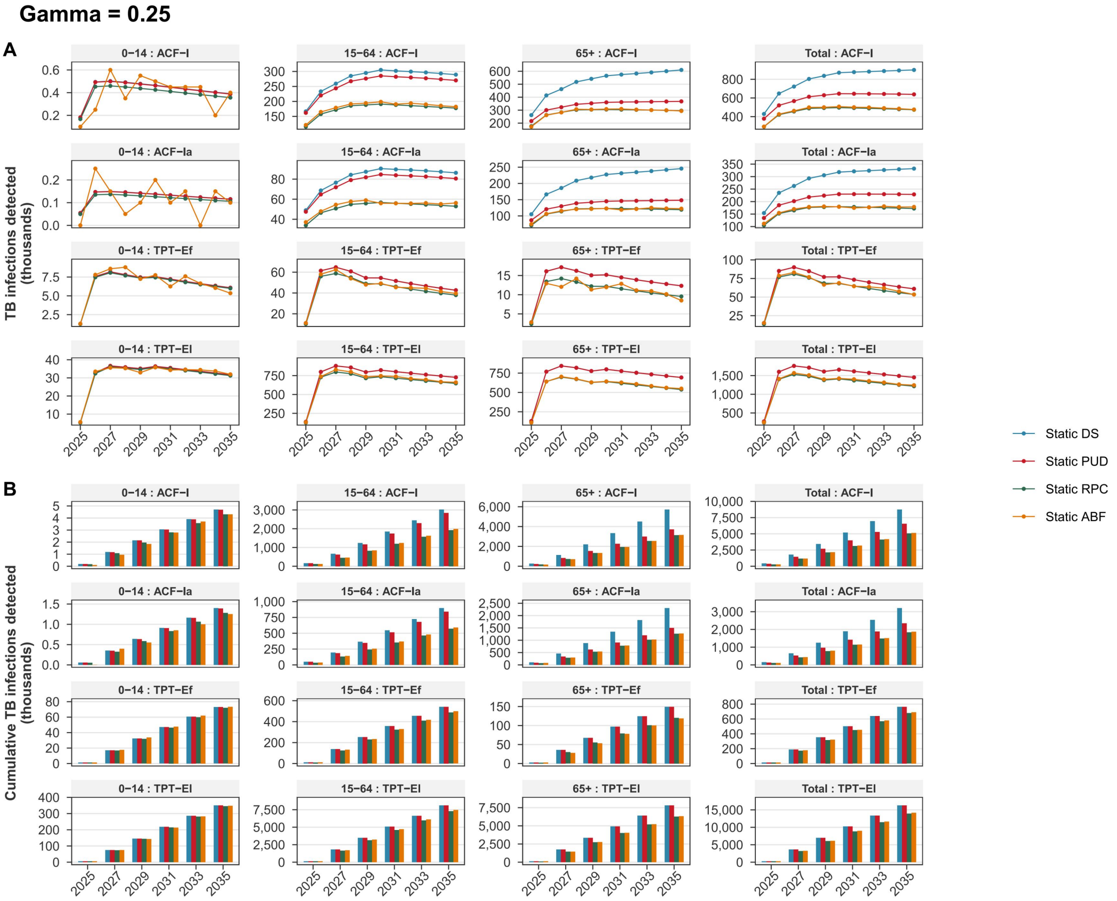
Gamma = 0.25
A
B
TB infections detected (thousands)
Cumulative TB infections detected (thousands)
0–14 : ACF–I
15–64 : ACF–I
65+ : ACF–I
Total : ACF–I
0–14 : ACF–Ia
15–64 : ACF–Ia
65+ : ACF–Ia
Total : ACF–Ia
0–14 : TPT–Ef
15–64 : TPT–Ef
65+ : TPT–Ef
Total : TPT–Ef
0–14 : TPT–El
15–64 : TPT–El
65+ : TPT–El
Total : TPT–El
2025
2027
2029
2031
2033
2035
Static DS
Static PUD
Static RPC
Static ABF

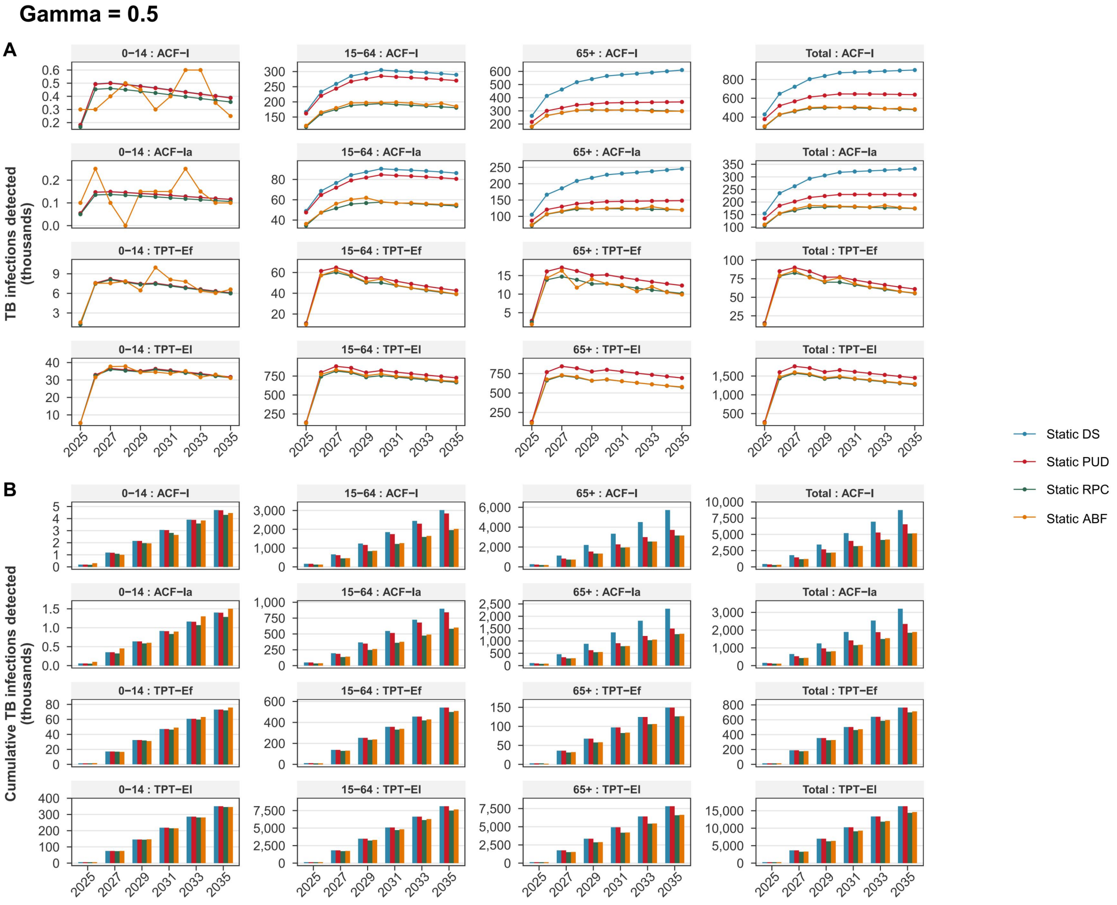
Gamma = 0.5
A
B
TB infections detected (thousands)
Cumulative TB infections detected (thousands)
0–14 : ACF–I
15–64 : ACF–I
65+ : ACF–I
Total : ACF–I
0–14 : ACF–Ia
15–64 : ACF–Ia
65+ : ACF–Ia
Total : ACF–Ia
0–14 : TPT–Ef
15–64 : TPT–Ef
65+ : TPT–Ef
Total : TPT–Ef
0–14 : TPT–El
15–64 : TPT–El
65+ : TPT–El
Total : TPT–El
2025
2027
2029
2031
2033
2035
Static DS
Static PUD
Static RPC
Static ABF

**Gamma = 1**

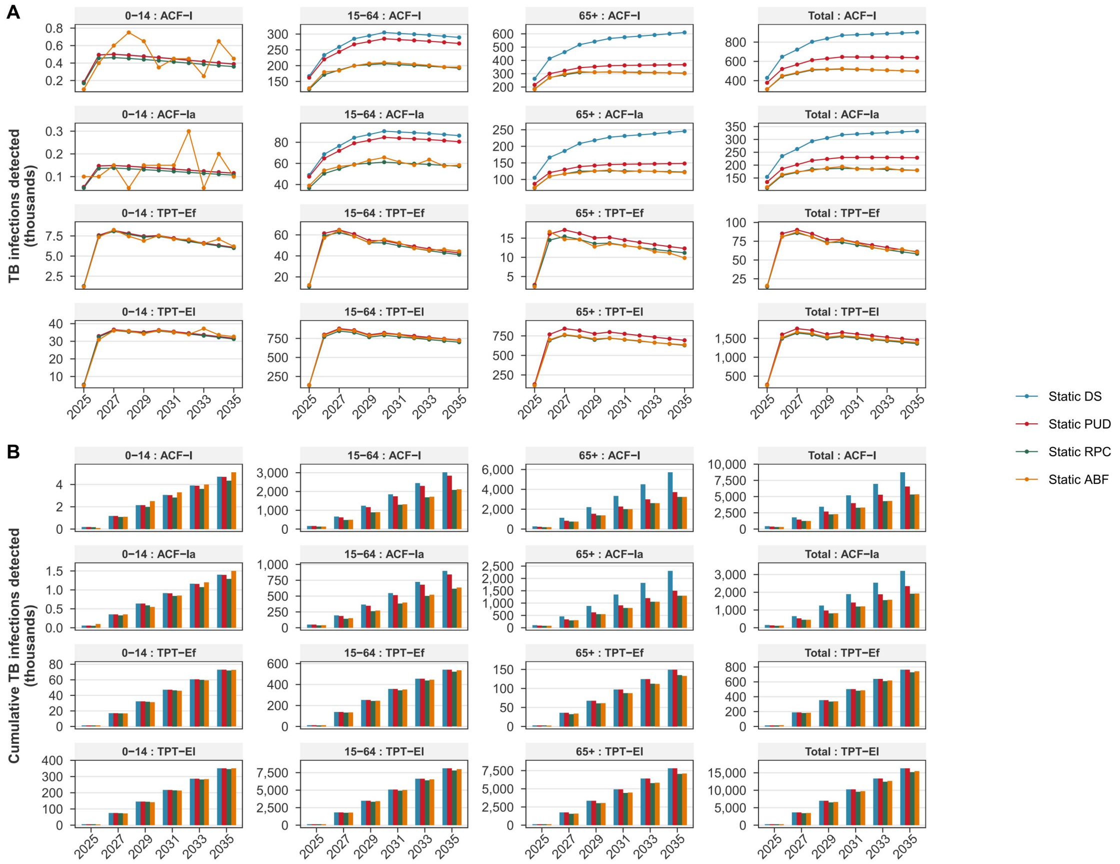


**Gamma = 2**

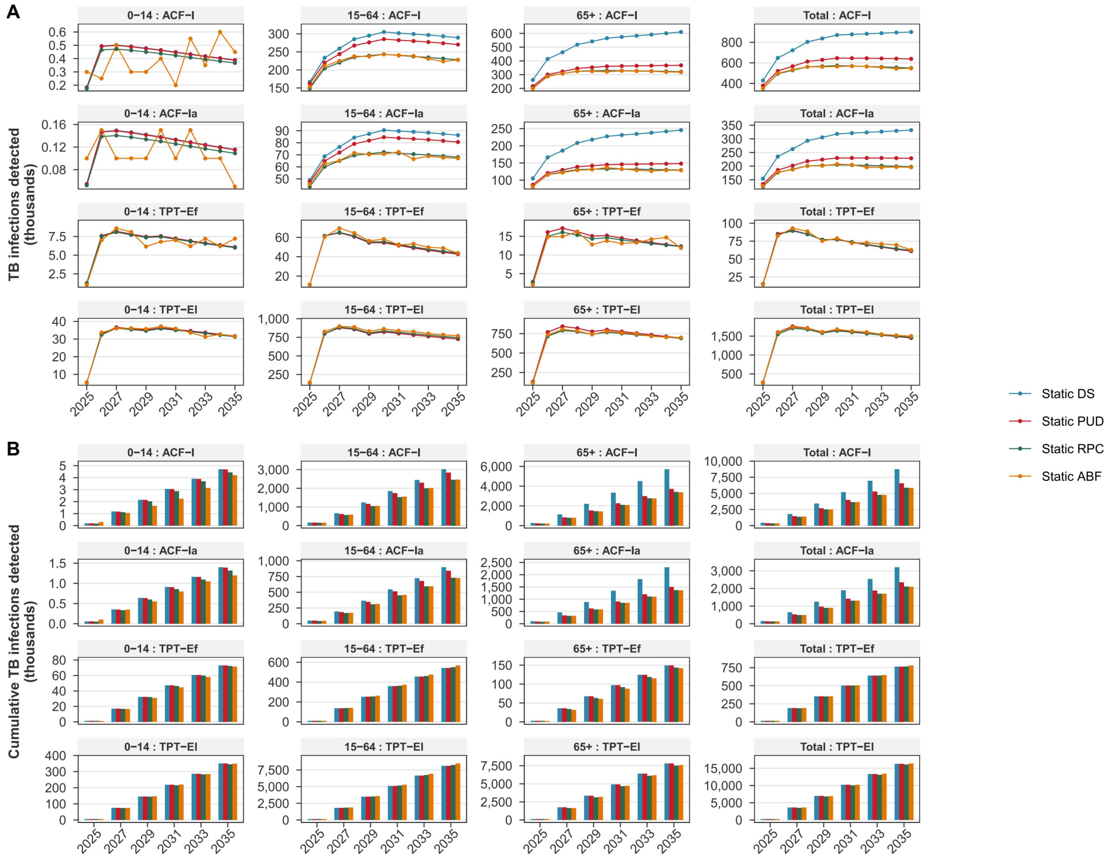

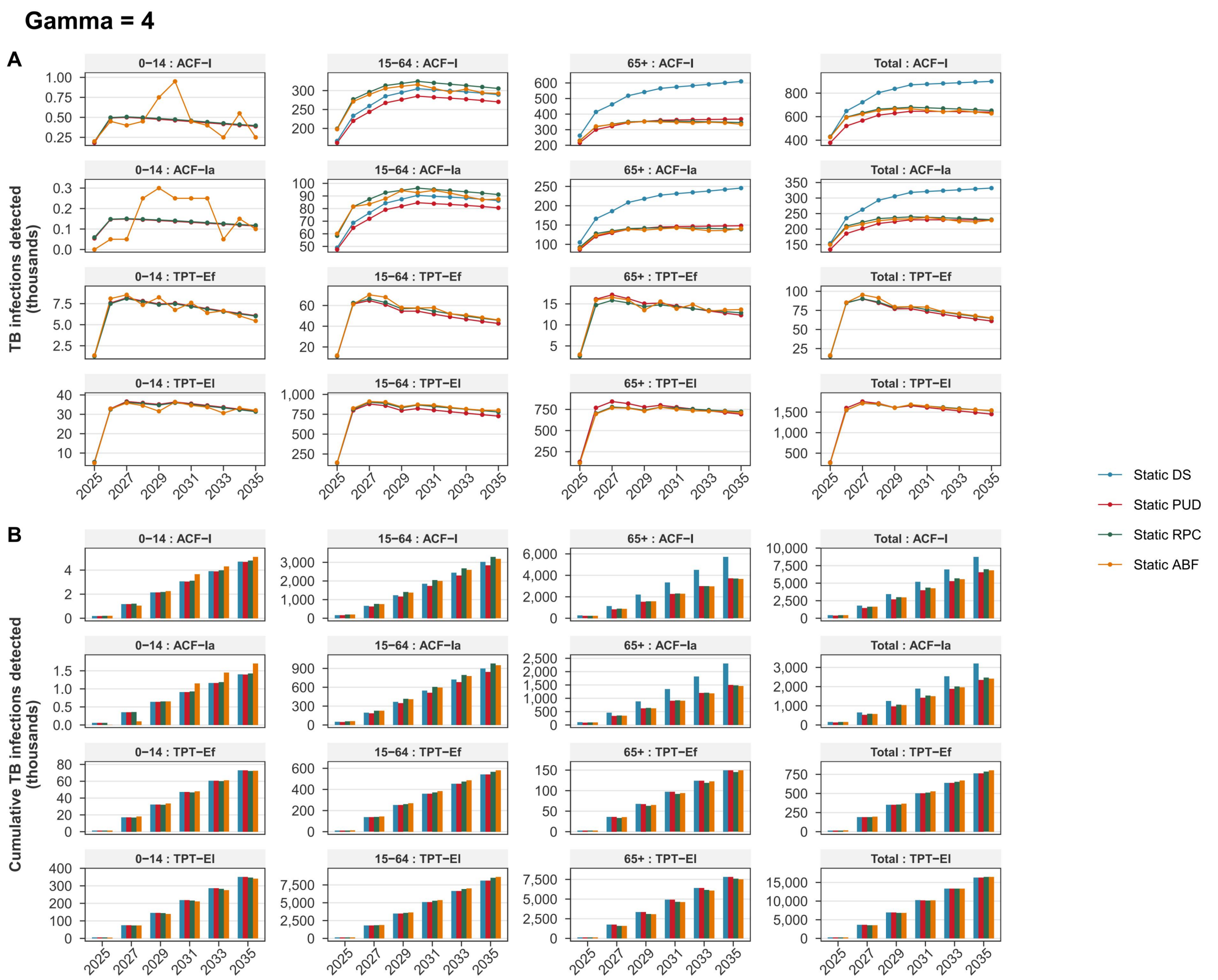


**Figure S6. Proportion of TB infection ($E_f$, $E_l$, $I$, $I_a$) detected by age group under different** $\gamma$ (0.25 to 4)**, 2025-2035.** $I_a$: asymptomatic active TB cases; $E_f$: recent LTBIs; $E_l$: remote LTBIs; $\gamma$, interaction parameter capturing the combined effects of overlapping risk factors.

**Table S1 TB transmission dynamic compartment model parameters and descriptions.**

| Parameter | Symbol | Value | Source |
|---|---|---|---|
| **TB natural history** | | | |
| Transmission coefficient | $\beta$ | - | To be calibrated |
| Scaling factor of the transmission ability of asymptomatic TB relative to symptomatic TB | $b$ | 0.7 | Ref.[3] |
| Protection from reinfection | $c$ | 0.5 | Assumption |
| Per-capita rate of progression from recent LTBIs to asymptomatic TB | $u$ | - | To be calibrated |
| Per-capita rate of stabilisation of recent LTBIs | $v$ | - | To be calibrated |
| Per-capita rate of reactivation from remote LTBIs to asymptomatic TB | $w$ | - | To be calibrated |
| Per-capita hazard of relapse, recently treated | $\rho_f$ | 0.048 $year^{(-1)}$ | WHO |
| Per-capita hazard of relapse, stabilised relapse risk | $\rho_l$ | 0.0015 $year^{(-1)}$ | WHO |
| Per-capita rate of stabilising relapse risk | $\phi$ | 1/2 $yr^{-1}$ | Duration conversion |
| **Age flow** | | | |
| Inflow from younger age group | $AgeIn$ | 1/15, 1/50 $year^{-1}$ | Duration conversion |
| Outflow to older age group | $AgeOut$ | 1/15, 1/50 $year^{-1}$ | Duration conversion |
| **Clinical course of TB disease** | | | |
| Per-capita mortality hazard, untreated TB | $\mu_{TB}$ | - | To be calibrated |
| Per-capita rate of treatment completion | $\tau$ | 2 $year^{-1}$ | Duration conversion |
| Progression rate from asymptomatic TB to symptomatic TB | $\theta_a$ | 2 $year^{-1}$ | Ref.[4] |
| Probability of self-healing for asymptomatic TB | $p_{sc}$ | 0.18 | Ref.[5] |
| Self-healing rate of Asymptomatic TB | $\theta_{sc}$ | 0.4 $year^{-1}$ | Ref.[5] |
| Self-healing rate of Symptomatic TB | $\theta$ | 1.72 $year^{-1}$ | Ref.[6] |
| **Other parameters** | | | |
| Recent LTBIs detecting rate through screening | $\sigma_f$ | To be Calculated (0 in the baseline scenario) | Multi-risk screening methods |

| | | | |
|---|---|---|---|
| remote LTBIs detecting rate through screening | $\sigma_l$ | To be Calculated (0 in the baseline scenario) | Multi-risk screening methods |
| Asymptomatic TB detecting rate through screening | $\sigma_a$ | To be Calculated (0 in the baseline scenario) | Multi-risk screening methods |
| Symptomatic TB detecting rate through screening | $\sigma_I$ | To be Calculated (0 in the baseline scenario) | Multi-risk screening methods |
| Preventive therapy acceptance rate * completion rate | $\alpha_p$ | 0.642 | Ref.[7] |
| Preventive therapy failure rate | $\gamma_p$ | 0.44 | Ref.[7, 8] |
| Preventive therapy implementation rate | $\theta_p$ | $4\ yr^{-1}$ | Duration conversion |
| Waning rate of Preventive therapy | $\theta_{pw}$ | $0.5\ yr^{-1}$ | Ref. [8, 9] |
| Preventive therapy efficacy | $P_E$ | 0.8 | Ref.[8] |
| Active case finding coverage of symptomatic TB | $p_a$ | 0.04 | TBIMS |
| Scaling factor of active case finding coverage of asymptomatic TB symptomatic TB | $p_b$ | 0.8 | TBIMS |
| Passive case finding coverage of symptomatic TB | $p_p$ | 0.75 | Assumption |
| Passive case detection rate | $\theta_{pa}$ | $13\ yr^{-1}$ | Duration conversion |
| Natural mortality rate | $\mu$ | Time dependent | Ref.[10] |

TB: Tuberculosis; WHO: World Health Organization.

**Table S2 Description and sources of data.**

| Data | Description | Source |
|---|---|---|
| Intervention scenarios | | |
| The baseline scenario | Scenario for maintaining 2024 TB screening coverage levels across various interventions from 2025 to 2035 | China CDC |
| The NTSP scenario | Scenario for TB screening coverage levels of the National Tuberculosis Strategic Plan (2025–2035) | China CDC |
| Population data | Age-stratified population size, crude birth rate and crude death rate from 2010 to 2023, with projections to 2035 | World Bank |
| Contact patterns | National-level contact matrix for 3×3 age groups | Ref.[11] |
| TB data | | |
| TB incidence | Number of incident TB cases nationwide for total population from 2010 to 2023 | WHO |
| TB mortality | Number of deaths caused by TB nationwide for total population from 2010 to 2023 | WHO |
| TB case notifications | Number of reported TB cases nationwide by age group from 2010 to 2023 | China CDC |
| High risk population | | |
| Close contact | Number of close contacts for active TB cases nationwide by age group from 2025 to 2035 | Compartment model |
| $HIV$ | Number of individuals living with HIV by age group in 2023, with projections to 2035 | China CDC |
| $HIV_{new}$ | Number of individuals newly diagnosed with HIV by age group in 2023, with projections to 2035 | China CDC |
| $R_f$ | Number of individuals recently recovered from active TB from 2025 to 2035 | Compartment model |
| $D_{new}$ | Number of individuals newly diagnosed with diabetes mellitus by age group in 2023, with projections to 2035 | Ref.[12] |
| $D_{pc}$ | Number of individuals living with diabetes mellitus and poor glycemic control by age group in 2023, with projections to 2035 | Ref.[12] |

TB: Tuberculosis; WHO: World Health Organization; $HIV$: individuals living with HIV; $HIV_{new}$: Individuals newly diagnosed with HIV; $R_f$: recently recovered individuals; 65+: Individuals aged 65 and above; $D_{new}$: Individuals newly diagnosed with diabetes mellitus; $D_{pc}$: Individuals living with diabetes mellitus and poor glycemic control.

**Table S3 TB infection rate multiplier (Multiplier 1).**

| Risk group | TB infection rate | Prevalence ratio* | Study design | Study period | Study site | Study population | Reference | Note |
|---|---|---|---|---|---|---|---|---|
| General population | 18.1% | _ | Cross-sectional study | 2013 | China | > 5 years old | Ref.[13] | |
| Individuals living with HIV | 14.4% | 0.8 | Meta analysis | 2000-2012 | China | 0-100 years old | Ref.[14] | |
| Individuals living with diabetes mellitus | - | 1.47 | Meta analysis | 2006-2021 | Global | 0-100 years old | Ref.[15] | Estimated from the Odds ratio: $PR = \frac{OR}{(1-P)+(P*OR)}$ |
| Close contact | 25.0% | 1.38 | Meta analysis | 2003-2022 | Global | 0-15 years | Ref.[16] | |
| Close contact | 24.0% | 1.33 | Meta analysis | 2003-2022 | Global | 15-60 years | Ref.[16] | |
| Close contact | 31.0% | 1.71 | Meta analysis | 2003-2022 | Global | 60+ years | Ref.[16] | |

*Prevalence ratio denotes the ratio of the TB infection rate in the risk factor group to that in the general population. HIV: individuals living with HIV; PR: Prevalence ratio; OR, odds ratio; P, prevalence.

**Table S4 Active TB prevalence multiplier among infected individuals (Multiplier 2).**

| Risk group | Active TB prevalence | Prevalence ratio* | Study design | Study period | Study site | Study population | Reference | Note |
|---|---|---|---|---|---|---|---|---|
| General population | 464/100,000 | _ | Cross-sectional study | 2013 | China | 0-100 years old | Ref.[17] | |
| Individuals recovered from TB within 2 years | 14,010/100,000 | 30.2 | Cross-sectional study | 2013 | China | 0-100 years old | Ref.[17] | |
| Individuals living with HIV | 7,200/100,000 | 15.5/0.8 | Meta analysis | 2000-2012 | China | 0-100 years old | Ref.[18] | |
| Individuals living with diabetes mellitus | - | 7.18/1.47 | Meta analysis | 1995-2015 | Global | 0-100 years old | Ref.[19] | Estimated from the Risk ratio: $PR = RR * \frac{T_{DM}}{T_{Non-DM}}$ |
| Close contact | - | 6.86/1.38 | Cohort study | 2001-2013 | China | 0-15 years | Ref.[20] | A substantial number of recent infections have been identified among close contacts, necessitating a high prevalence correction. |
| Close contact | - | 6.61/1.33 | Cohort study | 2001-2013 | China | 15-60 years | Ref.[20] | |
| Close contact | - | 8.50/1.71 | Cohort study | 2001-2013 | China | 60+ years | Ref.[20] | |

*Prevalence ratio denotes the ratio of the TB prevalence among infected individuals in the risk factor group to that among infected individuals in the general population. HIV: individuals living with HIV; PR: Prevalence ratio; RR, risk ratio; T, duration of illness.

**Table S5 Initial values of 9 epidemiological compartments by age group of 2010.**

| Symbol | Description | 0-14 | 15-64 | 65+ |
|---|---|---|---|---|
| $S$ | Susceptible population | 239,447,300 | 790,885,300 | 60,910,430 |
| $E_f$ | Recent LTBIs | 1,561,237 | 43,278,890 | 10,434,240 |
| $E_l$ | Remote LTBIs | 4,683,711 | 129,836,700 | 31,302,720 |
| $P$ | LTBIs protected by preventive therapy | 0 | 0 | 0 |
| $I_a$ | Asymptomatic active TB cases | 9,648 | 771,544 | 198,488 |
| $I$ | Symptomatic active TB cases | 14,472 | 1,388,779 | 406,900 |
| $T$ | TB cases under isolation and treatment | 4,367 | 349,221 | 116,793 |
| $R_f$ | Recently recovered individuals | 14,338 | 1,375,895 | 530,945 |
| $R_l$ | remotely recovered individuals | 129,040 | 12,383,060 | 4,778,512 |

TB: Tuberculosis; LTBI: Latent Tuberculosis Infection.
